# Progression of Carotid Intima-Media Thickness in Children of the Cardiovascular Comorbidity in Children with Chronic Kidney Disease Study (4C Study) – Risk Factors and Impact of Blood Pressure Dynamics

**DOI:** 10.1101/2024.07.13.24310158

**Authors:** Anke Doyon, Jonas Hofstetter, Aysun Karabay Bayazit, Karolis Azukaitis, Ana Niemirska, Mahmut Civilibal, Ipek Kaplan Bulut, Ali Duzova, Berna Oguz, Bruno Ranchin, Rukshana Shroff, Yelda Bilginer, Salim Caliskan, Dusan Paripovic, Cengiz Candan, Alev Yilmaz, Jerome Harambat, Z. Birsin Özçakar, Francesca Lugani, Harika Alpay, Sibylle Tschumi, Ebru Yilmaz, Dorota Drozdz, Yilmaz Tabel, Gül Özcelik, Alberto Caldas Afonso, Onder Yavascan, Anette Melk, Uwe Querfeld, Franz Schaefer, the 4C Study Consortium

## Abstract

**Rationale:** The progression of cardiovascular disease (CVD) in children with chronic kidney disease (CKD) is not well understood.

**Objective:** To investigate carotid intima-media thickness (cIMT) as a surrogate marker for CVD in 670 patients of the 4C Study (The Cardiovascular Comorbidity in Children with CKD Study), aged 6 - 17 years, with CKD stage 3-5 at baseline. Patients were observed for a period of up to 8 years.

**Methods and Results:** A linear mixed model was employed to analyse the longitudinal course of cIMT standard deviation score (SDS) and associated clinical risk factors. The association between cIMT SDS change rate and systolic and diastolic BP SDS change rate per year was investigated. cIMT SDS increased significantly during the prospective observation period, with the slope of increase attenuating over time. Younger, taller and female patients were at an elevated risk for elevated cIMT SDS. Further risk factors included diastolic blood pressure and serum albumin for all patients, albuminuria in progressive CKD, and serum phosphate in stable CKD. Diastolic blood pressure SDS decreased and its effect on cIMT SDS attenuated over time. The yearly diastolic and systolic blood pressure change rates were associated with the cIMT SDS change rate within the first 4.5 years. This indicates a progressive change in cIMT with increasing blood pressure over time, and a decrease in cIMT with lowering of blood pressure.

**Conclusions:** The results demonstrate a progressive increase in cIMT over time in children with CKD, with traditional risk factors such as albuminuria, serum phosphate and blood pressure as relevant predictive factors for cIMT SDS. The association of cIMT SDS progression with blood pressure dynamics suggests potential benefits of blood pressure control in children with CKD. Our findings indicate that cIMT may serve as a surrogate parameter for future clinical trials in children.

## Introduction

Cardiovascular disease (CVD) is a major complication of chronic kidney disease (CKD) and the most important risk factor for long-term patient survival. While subclinical signs of CVD already manifest in children and adolescents with CKD,^12^ the pace of deterioration and possible impact of risk factor modification is currently unclear. Prospective studies in children may be uniquely suited to reveal the intrinsic effects of CKD on the cardiovascular system due to the absence of comorbid conditions related to ageing, diabetes and smoking. To this end, the Cardiovascular Comorbidity in Children with CKD (4C) Study, initiated in 2009, prospectively followed 704 patients aged 6 to 17 years with a baseline glomerular filtration rate of 10 to 60 ml/min/1.73m^2^ in 55 pediatric nephrology units across 12 European countries. The morphology and function of the heart and the large arteries were regularly assessed using non-invasive methods including measurement of the carotid intima media thickness (cIMT), a validated surrogate marker of CVD commonly used for risk stratification^3^. Multiple clinical, anthropometric, biochemical and pharmacological risk factors were monitored prospectively and related to the cardiovascular status of the patients. Follow-up was continued when patients reached end-stage kidney disease (ESKD), and the effects of different kidney replacement modalities were addressed in separate sub-studies^4,5^.

Distinct alterations of the cardiovascular system were observed at the baseline characterization of the 4C cohort, including significant increases in cIMT, pulse wave velocity and left ventricular mass index and a high prevalence of uncontrolled hypertension^6^. Pulse wave velocity as a surrogate marker of arterial stiffness has been analyzed longitudinally and blood pressure was identified as a significant predictive factor.^7^ The present study complements these findings by a longitudinal analysis of cIMT progression over the course of 8 years in the overall cohort and two sub-cohorts, an assessment of risk factors associated with increased cIMT and the association between blood pressure dynamics and cIMT progression over time.

## Methods

### Measurement of Carotid Intima Media Thickness (cIMT) and of Covariates

cIMT was measured in patients of the 4C Study according to the Mannheim Intima-Media-Thickness consensus^8^. Measurements were performed with a mobile cardiovascular ultrasound device (Acuson P50, Siemens Medical Solutions USA, Inc.) and image analysis was carried out using Syngo US Workplace, Siemens Medical Solutions USA, Inc).

### Data Preprocessing & Inclusion of Patients and Visits

The 4C study was approved by the Ethics Committee of Heidelberg University (S-032/2009) and the institutional review boards at each participating institution^9^. Written informed consent was obtained from all parents and participants as appropriate. The study was registered at ClinicalTrials.gov August 7, 2009, Identifier NCT01046448).

In the 4C Study, regional investigators were trained and travelled yearly to individually assigned sites to perform cardiovascular measurements (pulse wave velocity, cIMT recording, echocardiography, blood pressure measurements) in participants. At the yearly visit and at additional 6-monthly visits, patient history and anthropometry were recorded and laboratory work-up was performed and analyzed both locally and centrally. All patients with at least one valid cIMT measurement before dropout or before initiation of kidney replacement therapy (KRT) and with eGFR <60 ml/min/1.73m^2^ were included in the analysis. cIMT measurements took place at the annual clinical visits or close to this date (maximally ± 3 months). All visits until 8 years after the baseline visit were included. Visits after transplantation or initiation of dialysis were not taken into consideration. This resulted in inclusion of 670 patients with at least one valid cIMT measurement and a total of 2,221 visits with valid cIMT values. The first recording was used as individual baseline for further analysis in the multivariable longitudinal model if follow-up data were available. (Figure 1).

**Figure 1.**
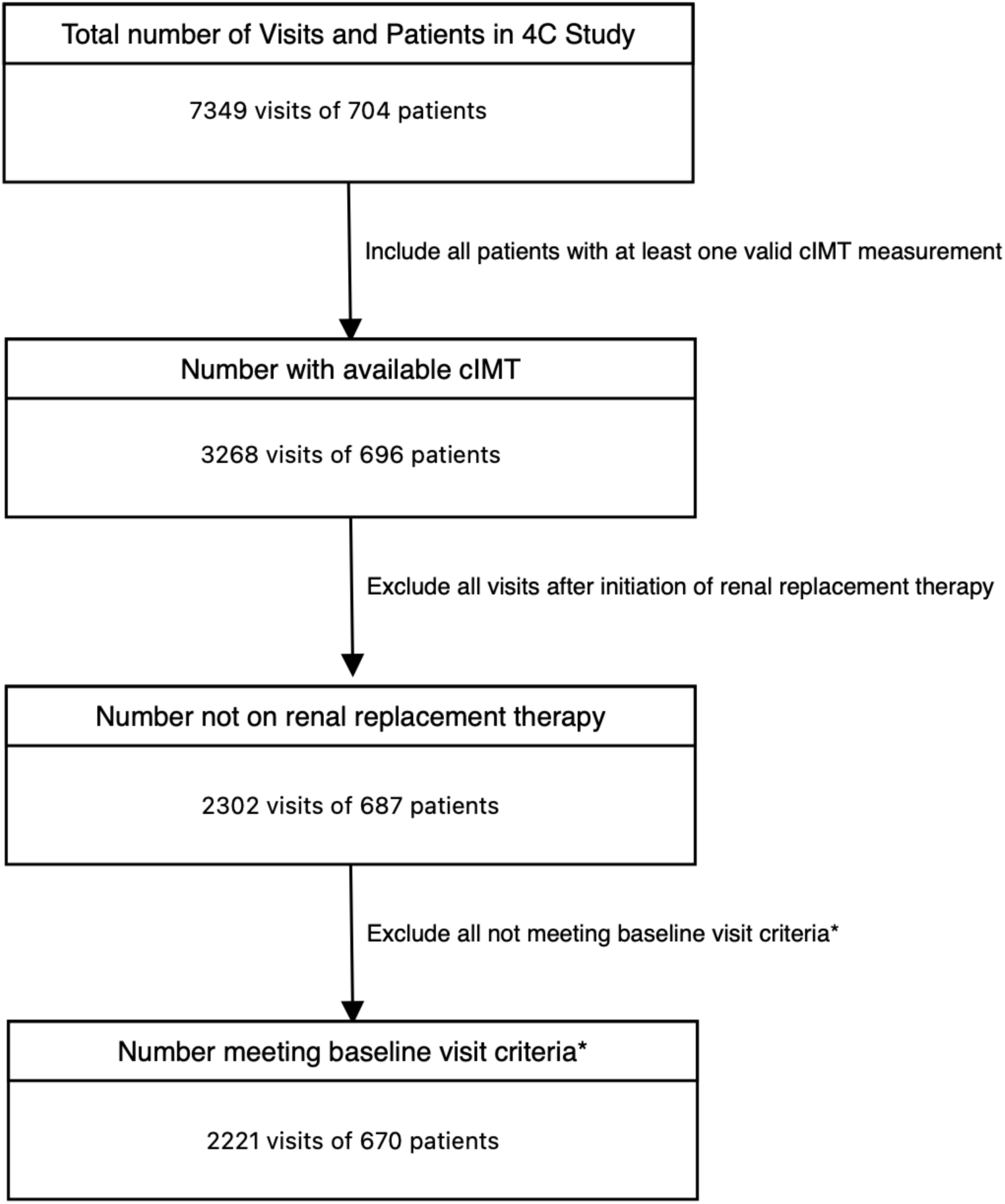
Patient and visit selection (*baseline visit criteria: eGFR<60 ml/min/1.73m^2^, age 6-17 years)

The measured cIMT values were standardized for sex and age by calculation of standard deviation scores (SDS; normal range +2 to −2) as described previously^8^. Biochemical analyses were performed centrally except for hemoglobin, intact parathyroid hormone (iPTH), serum bicarbonate and ferritin. Standard laboratory techniques were used to measure the serum and urinary concentrations of albumin and creatinine and the serum concentrations of phosphorus, calcium, albumin, iPTH, bicarbonate, LDL and HDL cholesterol, C-reactive protein (CRP) and uric acid. The estimated glomerular filtration rate (eGFR) was calculated according to the updated Schwartz equation^10^. Blood pressure (BP) was measured locally with oscillometric devices validated for pediatric use and appropriate cuff sizes. BP values were normalized to SDS according to age, sex and height^11^. Height and BMI SDS were calculated based on WHO reference data.

### Descriptive Statistics

Descriptive statistics of cIMT SDS, anthropometric measures, CKD stage, biochemical measures, and antihypertensive therapy status were calculated for the annual visits across all patients. Stratification was performed for stable eGFR vs. progressive kidney failure defined by the attainment of a composite endpoint including 50% eGFR loss, eGFR below 10 ml/min/1.73m^2^, or start of KRT. Descriptive statistics were calculated as counts (percentage) for categorical measures, mean (SD) for continuous measures with normal distribution and median (interquartile range) for those with skewed distribution.

A linear mixed model (LMM) was formulated that could make inference to time-varying, average population-level relationships between explanatory covariates and cIMT SDS. The modeling approach aimed at flexibly incorporating properties of the risk factor relationships with cIMT SDS discovered during visual data exploration, and at remedying for LMM assumption violations evoked by particularities in the longitudinal data set (see supplemental material for details).

### An Application-Specific Linear Mixed Model Formulation

Separate LMM were specified for longitudinal modeling of cIMT SDS in all patients, in patients with progressive chronic kidney disease (P-CKD), and in patients with stable chronic kidney disease (S-CKD). The detailed development of the models and related diagnostics are presented in the supplements^12^.

The final LMM per patient group (all patients, patients with P-CKD, patients with S-CKD) was fitted to 20 imputed datasets and the estimated LMM parameters were pooled according to Rubin’s Rule^13^. The pooled fixed effect estimates, the pooled center-level and patient-level random effect covariance parameters, the derived cIMT SDS variability contributions at each hierarchical level of the model structure (average population-level, center-level, patient-level), and the derived *intra-patient-level* correlations for specific time intervals of follow-up are reported in the results section and the supplement of this paper for the final LMM per patient group. The lme function from the nlme package^14^ was utilized in R Studio vs 4.2.2^15,16^ for longitudinal linear mixed modeling.

### Post-fit correlations of absolute linear patient-level change per year in cIMT SDS with absolute linear patient-level change per year in blood pressure SDS

The LMM model set up for all patients as described above was then used to obtain yearly rates of absolute changes for cIMT SDS (δ cIMT SDS), systolic BP SDS (δ systolic BP SDS) and diastolic BP SDS (δ diastolic BP SDS). δ cIMT SDS for each patient was calculated from predicted values of the elaborated LMM and δ systolic BP SDS and δ diastolic BP SDS were derived directly from the estimated predictive parameters of the LMM. For this analysis, data until a maximum follow-up of 4.5 years were taken into account (see supplemental methods for details). Subsequently, the linear relationship between δ cIMT BP SDS and either δ systolic BP SDS or δ diastolic BP SDS was modeled by linear regression analysis.

## Results

Figure 1 shows patient and visit selection according to the inclusion criteria. The majority of patients was diagnosed with CAKUT as underlying kidney disease (69%), 12.4% had a tubulointerstitial kidney disorder, 8.8% had a glomerulopathy, and 5.2 % had CKD following acute kidney injury (AKI) for various reasons (e.g. hemolytic uremic syndrome, sepsis, postnatal asphyxia). The percentage of patients with blood pressure medication increased at the 2-year and 4-year follow-up visit compared to baseline. Both systolic and diastolic blood pressure decreased during the course of the study (**Table 1**).

**Table 1:**
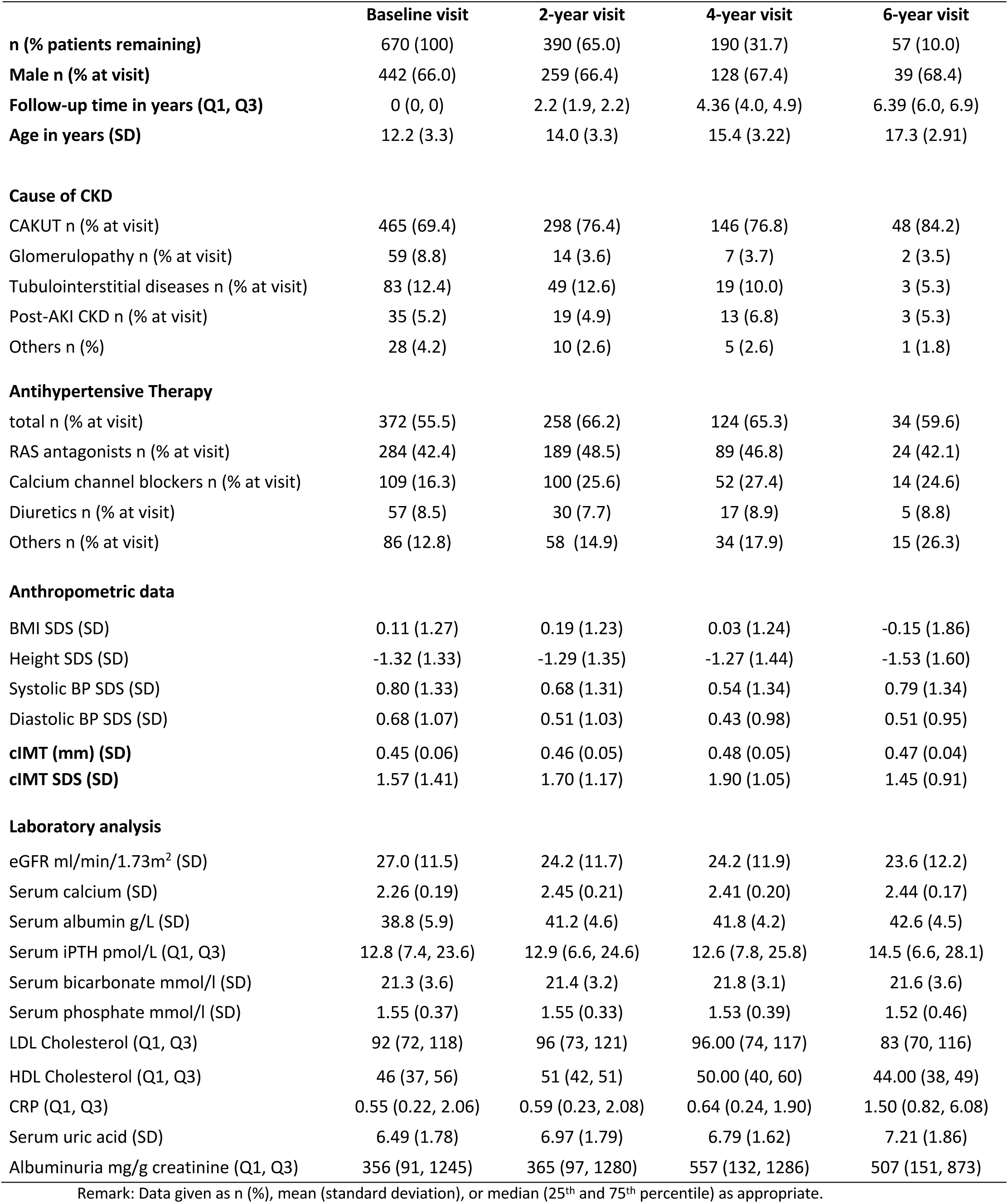
Patient Characteristics, according to visit.

|  | Baseline visit | 2-year visit | 4-year visit | 6-year visit |
| --- | --- | --- | --- | --- |
| **n (% patients remaining)** | 670 (100) | 390 (65.0) | 190 (31.7) | 57 (10.0) |
| **Male n (% at visit)** | 442 (66.0) | 259 (66.4) | 128 (67.4) | 39 (68.4) |
| **Follow-up time in years (Q1, Q3)** | 0 (0, 0) | 2.2 (1.9, 2.2) | 4.36 (4.0, 4.9) | 6.39 (6.0, 6.9) |
| **Age in years (SD)** | 12.2 (3.3) | 14.0 (3.3) | 15.4 (3.22) | 17.3 (2.91) |
| **Cause of CKD** |  |  |  |  |
| CAKUT n (% at visit) | 465 (69.4) | 298 (76.4) | 146 (76.8) | 48 (84.2) |
| Glomerulopathy n (% at visit) | 59 (8.8) | 14 (3.6) | 7 (3.7) | 2 (3.5) |
| Tubulointerstitial diseases n (% at visit) | 83 (12.4) | 49 (12.6) | 19 (10.0) | 3 (5.3) |
| Post-AKI CKD n (% at visit) | 35 (5.2) | 19 (4.9) | 13 (6.8) | 3 (5.3) |
| Others n (%) | 28 (4.2) | 10 (2.6) | 5 (2.6) | 1 (1.8) |
| **Antihypertensive Therapy** |  |  |  |  |
| total n (% at visit) | 372 (55.5) | 258 (66.2) | 124 (65.3) | 34 (59.6) |
| RAS antagonists n (% at visit) | 284 (42.4) | 189 (48.5) | 89 (46.8) | 24 (42.1) |
| Calcium channel blockers n (% at visit) | 109 (16.3) | 100 (25.6) | 52 (27.4) | 14 (24.6) |
| Diuretics n (% at visit) | 57 (8.5) | 30 (7.7) | 17 (8.9) | 5 (8.8) |
| Others n (% at visit) | 86 (12.8) | 58 (14.9) | 34 (17.9) | 15 (26.3) |
| **Anthropometric data** |  |  |  |  |
| BMI SDS (SD) | 0.11 (1.27) | 0.19 (1.23) | 0.03 (1.24) | -0.15 (1.86) |
| Height SDS (SD) | -1.32 (1.33) | -1.29 (1.35) | -1.27 (1.44) | -1.53 (1.60) |
| Systolic BP SDS (SD) | 0.80 (1.33) | 0.68 (1.31) | 0.54 (1.34) | 0.79 (1.34) |
| Diastolic BP SDS (SD) | 0.68 (1.07) | 0.51 (1.03) | 0.43 (0.98) | 0.51 (0.95) |
| **cIMT (mm) (SD)** | 0.45 (0.06) | 0.46 (0.05) | 0.48 (0.05) | 0.47 (0.04) |
| **cIMT SDS (SD)** | 1.57 (1.41) | 1.70 (1.17) | 1.90 (1.05) | 1.45 (0.91) |
| **Laboratory analysis** |  |  |  |  |
| eGFR ml/min/1.73m<sup>2</sup> (SD) | 27.0 (11.5) | 24.2 (11.7) | 24.2 (11.9) | 23.6 (12.2) |
| Serum calcium (SD) | 2.26 (0.19) | 2.45 (0.21) | 2.41 (0.20) | 2.44 (0.17) |
| Serum albumin g/L (SD) | 38.8 (5.9) | 41.2 (4.6) | 41.8 (4.2) | 42.6 (4.5) |
| Serum iPTH pmol/L (Q1, Q3) | 12.8 (7.4, 23.6) | 12.9 (6.6, 24.6) | 12.6 (7.8, 25.8) | 14.5 (6.6, 28.1) |
| Serum bicarbonate mmol/l (SD) | 21.3 (3.6) | 21.4 (3.2) | 21.8 (3.1) | 21.6 (3.6) |
| Serum phosphate mmol/l (SD) | 1.55 (0.37) | 1.55 (0.33) | 1.53 (0.39) | 1.52 (0.46) |
| LDL Cholesterol (Q1, Q3) | 92 (72, 118) | 96 (73, 121) | 96.00 (74, 117) | 83 (70, 116) |
| HDL Cholesterol (Q1, Q3) | 46 (37, 56) | 51 (42, 51) | 50.00 (40, 60) | 44.00 (38, 49) |
| CRP (Q1, Q3) | 0.55 (0.22, 2.06) | 0.59 (0.23, 2.08) | 0.64 (0.24, 1.90) | 1.50 (0.82, 6.08) |
| Serum uric acid (SD) | 6.49 (1.78) | 6.97 (1.79) | 6.79 (1.62) | 7.21 (1.86) |
| Albuminuria mg/g creatinine (Q1, Q3) | 356 (91, 1245) | 365 (97, 1280) | 557 (132, 1286) | 507 (151, 873) |

Remark: Data given as n (%), mean (standard deviation), or median (25<sup>th</sup> and 75<sup>th</sup> percentile) as appropriate.

Patient characteristics at baseline and at the 2-, 4- and 6-year follow up visits are given in **Table 1**., baseline characteristics according to progressive kidney disease or stable chronic kidney disease are displayed in **Table 2**. There were a median of 3 cIMT measurements per patient in both groups. The mean cIMT at baseline was 0.45 ± 0.06 mm, and the mean cIMT SDS was 1.57 ± 1.41. The cIMT was increased by more than 2 SDS in 32.4% of patients at study entry.

**Table 2:**
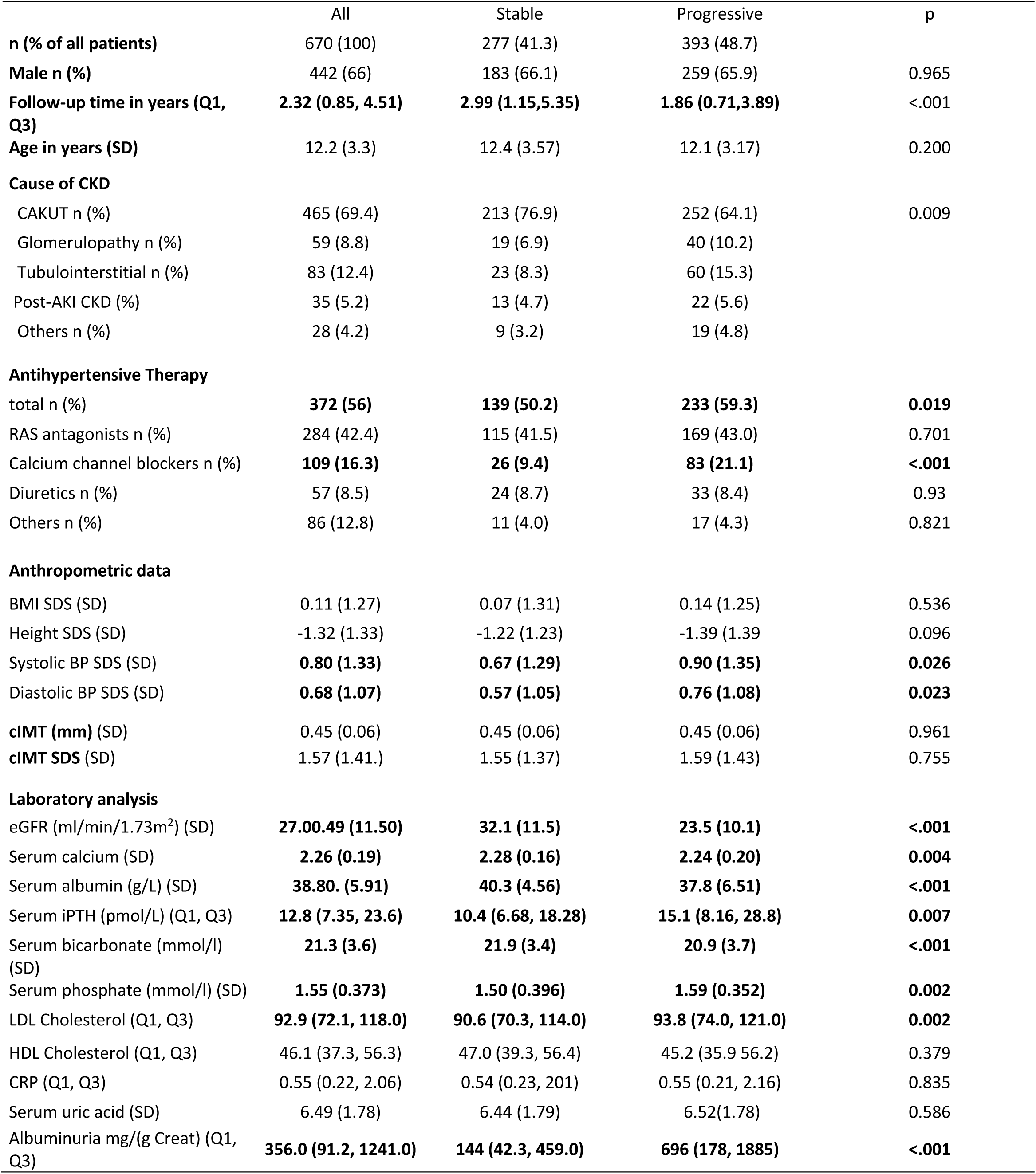
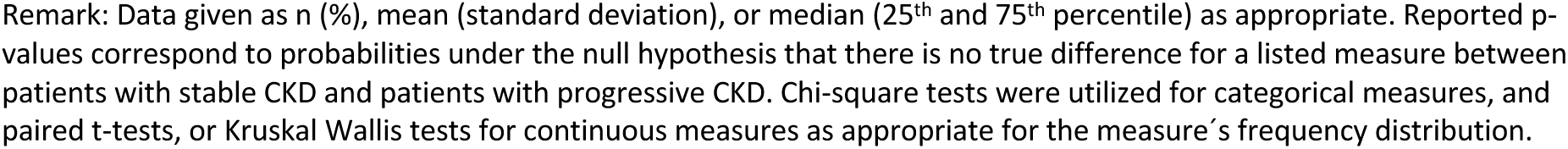
Characteristics of patients with stable CKD and of patients with progressive CKD at baseline.

|  | All | Stable | Progressive | p |
| --- | --- | --- | --- | --- |
| **n (% of all patients)** | 670 (100) | 277 (41.3) | 393 (48.7) |  |
| **Male n (%)** | 442 (66) | 183 (66.1) | 259 (65.9) | 0.965 |
| **Follow-up time in years (Q1, Q3)** | **2.32 (0.85, 4.51)** | **2.99 (1.15,5.35)** | **1.86 (0.71,3.89)** | <.001 |
| **Age in years (SD)** | 12.2 (3.3) | 12.4 (3.57) | 12.1 (3.17) | 0.200 |
| **Cause of CKD** |  |  |  |  |
| CAKUT n (%) | 465 (69.4) | 213 (76.9) | 252 (64.1) | 0.009 |
| Glomerulopathy n (%) | 59 (8.8) | 19 (6.9) | 40 (10.2) |  |
| Tubulointerstitial n (%) | 83 (12.4) | 23 (8.3) | 60 (15.3) |  |
| Post-AKI CKD (%) | 35 (5.2) | 13 (4.7) | 22 (5.6) |  |
| Others n (%) | 28 (4.2) | 9 (3.2) | 19 (4.8) |  |
| **Antihypertensive Therapy** |  |  |  |  |
| total n (%) | **372 (56)** | **139 (50.2)** | **233 (59.3)** | **0.019** |
| RAS antagonists n (%) | 284 (42.4) | 115 (41.5) | 169 (43.0) | 0.701 |
| Calcium channel blockers n (%) | **109 (16.3)** | **26 (9.4)** | **83 (21.1)** | **<.001** |
| Diuretics n (%) | 57 (8.5) | 24 (8.7) | 33 (8.4) | 0.93 |
| Others n (%) | 86 (12.8) | 11 (4.0) | 17 (4.3) | 0.821 |
| **Anthropometric data** |  |  |  |  |
| BMI SDS (SD) | 0.11 (1.27) | 0.07 (1.31) | 0.14 (1.25) | 0.536 |
| Height SDS (SD) | -1.32 (1.33) | -1.22 (1.23) | -1.39 (1.39) | 0.096 |
| Systolic BP SDS (SD) | **0.80 (1.33)** | **0.67 (1.29)** | **0.90 (1.35)** | **0.026** |
| Diastolic BP SDS (SD) | **0.68 (1.07)** | **0.57 (1.05)** | **0.76 (1.08)** | **0.023** |
| cIMT (mm) (SD) | 0.45 (0.06) | 0.45 (0.06) | 0.45 (0.06) | 0.961 |
| cIMT SDS (SD) | 1.57 (1.41.) | 1.55 (1.37) | 1.59 (1.43) | 0.755 |
| **Laboratory analysis** |  |  |  |  |
| eGFR (ml/min/1.73m<sup>2</sup>) (SD) | **27.00.49 (11.50)** | **32.1 (11.5)** | **23.5 (10.1)** | **<.001** |
| Serum calcium (SD) | **2.26 (0.19)** | **2.28 (0.16)** | **2.24 (0.20)** | **0.004** |
| Serum albumin (g/L) (SD) | **38.80. (5.91)** | **40.3 (4.56)** | **37.8 (6.51)** | **<.001** |
| Serum iPTH (pmol/L) (Q1, Q3) | **12.8 (7.35, 23.6)** | **10.4 (6.68, 18.28)** | **15.1 (8.16, 28.8)** | **0.007** |
| Serum bicarbonate (mmol/l) (SD) | **21.3 (3.6)** | **21.9 (3.4)** | **20.9 (3.7)** | **<.001** |
| Serum phosphate (mmol/l) (SD) | **1.55 (0.373)** | **1.50 (0.396)** | **1.59 (0.352)** | **0.002** |
| LDL Cholesterol (Q1, Q3) | **92.9 (72.1, 118.0)** | **90.6 (70.3, 114.0)** | **93.8 (74.0, 121.0)** | **0.002** |
| HDL Cholesterol (Q1, Q3) | 46.1 (37.3, 56.3) | 47.0 (39.3, 56.4) | 45.2 (35.9 56.2) | 0.379 |
| CRP (Q1, Q3) | 0.55 (0.22, 2.06) | 0.54 (0.23, 201) | 0.55 (0.21, 2.16) | 0.835 |
| Serum uric acid (SD) | 6.49 (1.78) | 6.44 (1.79) | 6.52(1.78) | 0.586 |
| Albuminuria mg/(g Creat) (Q1, Q3) | **356.0 (91.2, 1241.0)** | **144 (42.3, 459.0)** | **696 (178, 1885)** | **<.001** |

Remark: Data given as n (%), mean (standard deviation), or median (25<sup>th</sup> and 75<sup>th</sup> percentile) as appropriate. Reported p-values correspond to probabilities under the null hypothesis that there is no true difference for a listed measure between patients with stable CKD and patients with progressive CKD. Chi-square tests were utilized for categorical measures, and paired t-tests, or Kruskal Wallis tests for continuous measures as appropriate for the measure's frequency distribution.

A total of 393 patients suffered from progressive kidney failure (P-CKD). Of these, 161 patients lost >50% eGFR, 76 patients attained eGFR <10 ml/min/1.73m^2^ and 156 commenced KRT. All other patients (n=277) were considered having “stable” CKD (S-CKD). The two groups differed significantly for their eGFR at baseline (31.5 ± 12 vs. 20.9 ± 9.6 ml/min/1.73m^2^, p<.001), systolic and diastolic blood pressure (p=0.026/0.023), serum calcium (p=0.004), albumin (p<.001), serum phosphate (p=0.002), iPTH (p=0.007) and bicarbonate (p<.001) and albuminuria (p<.001). (**Table 2).** At baseline, the distribution of cIMT and cIMT SDS was virtually identical in patients with stable CKD and those with progressive CKD (**Table 2**).

Linear mixed modeling showed a significant positive linear effect (ß=0.20, p<0.0001) and a negative quadratic effect of time on cIMT (−0.02, p<0.0001), reflecting a slowing increase of cIMT towards the end of the observation period. This pattern of association was also detected in the subgroups with stable and progressive CKD **(Table 3)**.

**Table 3.**
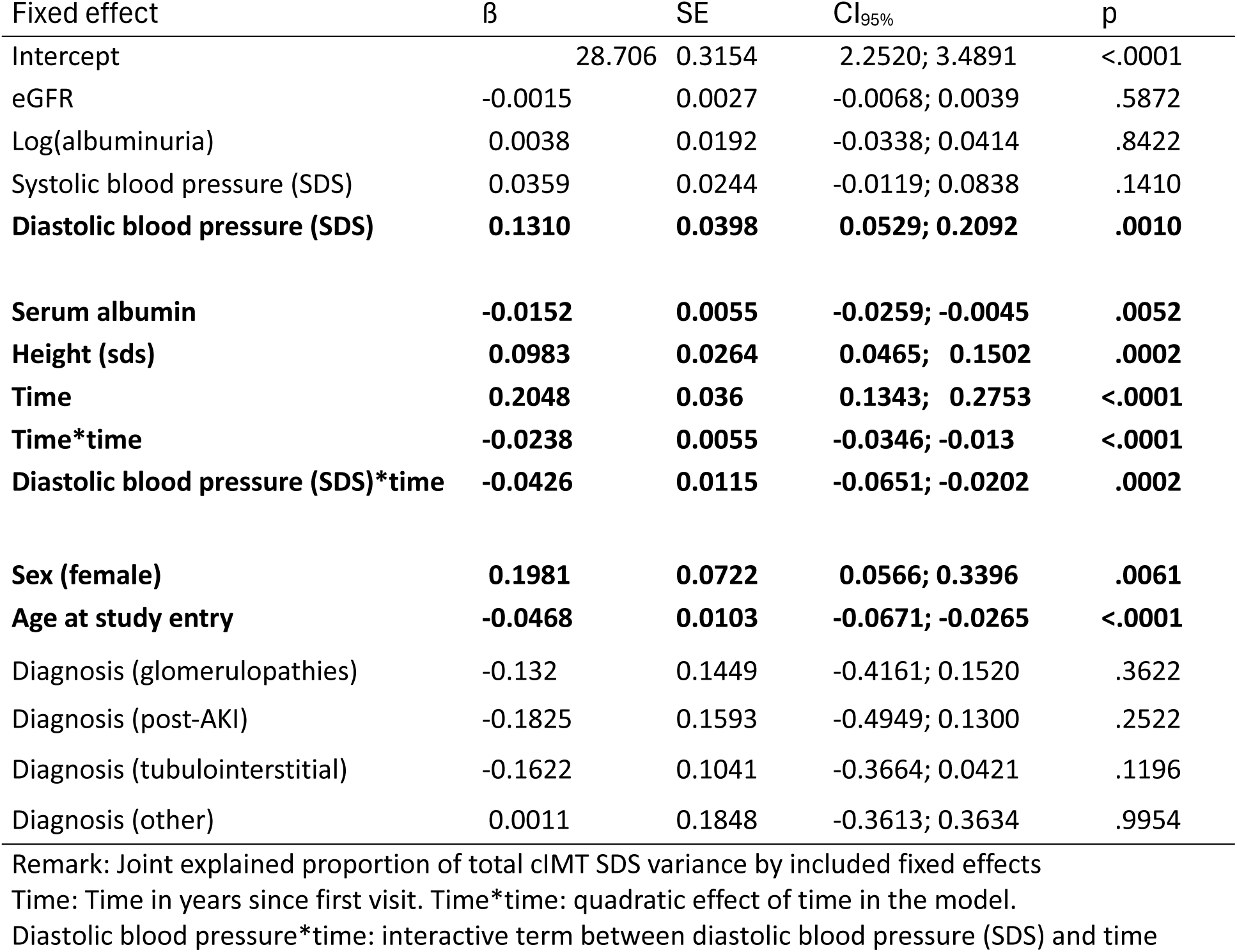
Pooled Population-Level (Fixed Effect) Mean Estimates of the Linear Mixed Model (LMM) with Modeled Time-Dependent Residual Variance Function for cIMT SDS for all Patients.

| Fixed effect | $\beta$ | SE | CI<sub>95%</sub> | p |
| --- | --- | --- | --- | --- |
| Intercept | 28.706 | 0.3154 | 2.2520; 3.4891 | <.0001 |
| eGFR | -0.0015 | 0.0027 | -0.0068; 0.0039 | .5872 |
| Log(albuminuria) | 0.0038 | 0.0192 | -0.0338; 0.0414 | .8422 |
| Systolic blood pressure (SDS) | 0.0359 | 0.0244 | -0.0119; 0.0838 | .1410 |
| **Diastolic blood pressure (SDS)** | **0.1310** | **0.0398** | **0.0529; 0.2092** | **.0010** |
| **Serum albumin** | **-0.0152** | **0.0055** | **-0.0259; -0.0045** | **.0052** |
| **Height (sds)** | **0.0983** | **0.0264** | **0.0465; 0.1502** | **.0002** |
| **Time** | **0.2048** | **0.036** | **0.1343; 0.2753** | **<.0001** |
| **Time\*time** | **-0.0238** | **0.0055** | **-0.0346; -0.013** | **<.0001** |
| **Diastolic blood pressure (SDS)\*time** | **-0.0426** | **0.0115** | **-0.0651; -0.0202** | **.0002** |
| **Sex (female)** | **0.1981** | **0.0722** | **0.0566; 0.3396** | **.0061** |
| **Age at study entry** | **-0.0468** | **0.0103** | **-0.0671; -0.0265** | **<.0001** |
| Diagnosis (glomerulopathies) | -0.132 | 0.1449 | -0.4161; 0.1520 | .3622 |
| Diagnosis (post-AKI) | -0.1825 | 0.1593 | -0.4949; 0.1300 | .2522 |
| Diagnosis (tubulointerstitial) | -0.1622 | 0.1041 | -0.3664; 0.0421 | .1196 |
| Diagnosis (other) | 0.0011 | 0.1848 | -0.3613; 0.3634 | .9954 |

Remark: Joint explained proportion of total cIMT SDS variance by included fixed effects
Time: Time in years since first visit. Time\*time: quadratic effect of time in the model.
Diastolic blood pressure\*time: interactive term between diastolic blood pressure (SDS) and time

In the total cohort, girls, younger and taller patients were at risk for higher cIMT SDS (ß=0.20, p=0.061; ß= −0.05, p<0.0001; and ß=0.10, p=.0002). Diastolic blood pressure was significantly predictive for cIMT SDS, while systolic BP showed only a borderline association (ß=0.036, p=0.141). Lower serum albumin levels were also associated with higher cIMT SDS (ß=-0.015, p<0.0052). The effects of age, height, sex and albumin remained stable throughout the study period. The negative interaction between diastolic BP and time (ß=-0.043, p=.001) indicated a decrease in the positive linear relationship between diastolic BP and cIMT SDS towards the end of the observation period **(Table 3)**.

In the group of patients with progressive CKD, albuminuria (ß=0.05, p=0.06) and diastolic blood pressure (ß =0.11, p=0.04) contributed to higher cIMT SDS, in addition to young age (ß=- 0.05,p=.0001), height SDS (ß=0.105, p=.0013), and female gender (ß=0.30, p=.0015) (**Table 5**). The effect of diastolic BP on cIMT SDS diminished significantly over time (ß=-0.06, p=.0002). In the stable CKD subgroup serum phosphate (ß=0.24, p=0.03) was the only significant risk factor for higher cIMT SDS in addition to time (**Table 4**). Figures 2-4 show sensitivity analyses of average cIMT SDS profiles as predicted by the fitted LMM for fixed median baseline values of the individual explanatory covariates and varied baseline values (at median ± 2*SD) of one selected explanatory covariate at baseline. Sensitivity plots in Figures 2-4 are provided separately for boys and girls as well as for the whole population,and the stable and progressive CKD subgroups, respectively.

**Figure 2.**
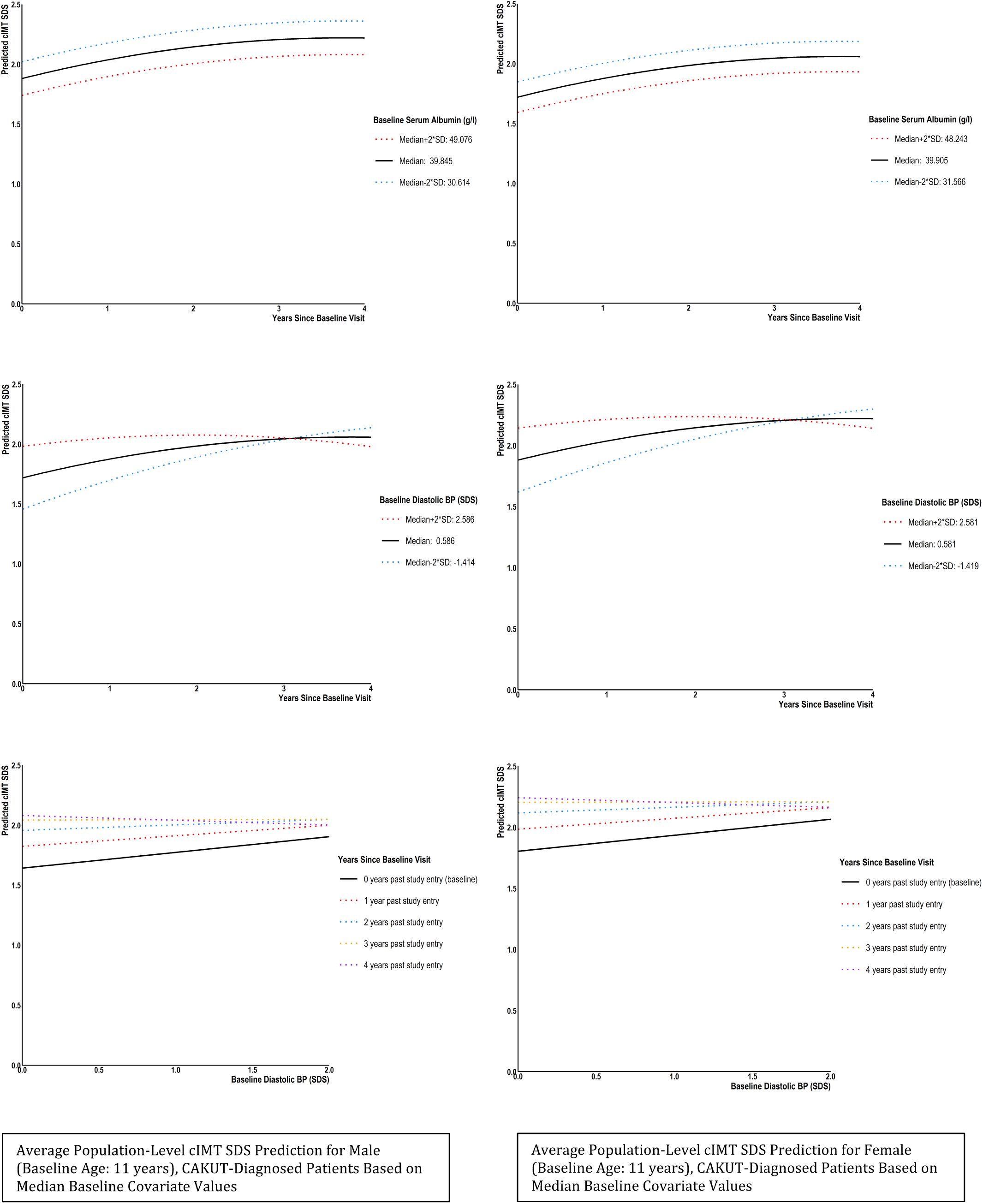
Average Population-Level cIMT SDS Prediction for Male and Female (Baseline Age: 11 years), CAKUT-Diagnosed Patients Based on Median Baseline Covariate Values.

**Figure 3.**
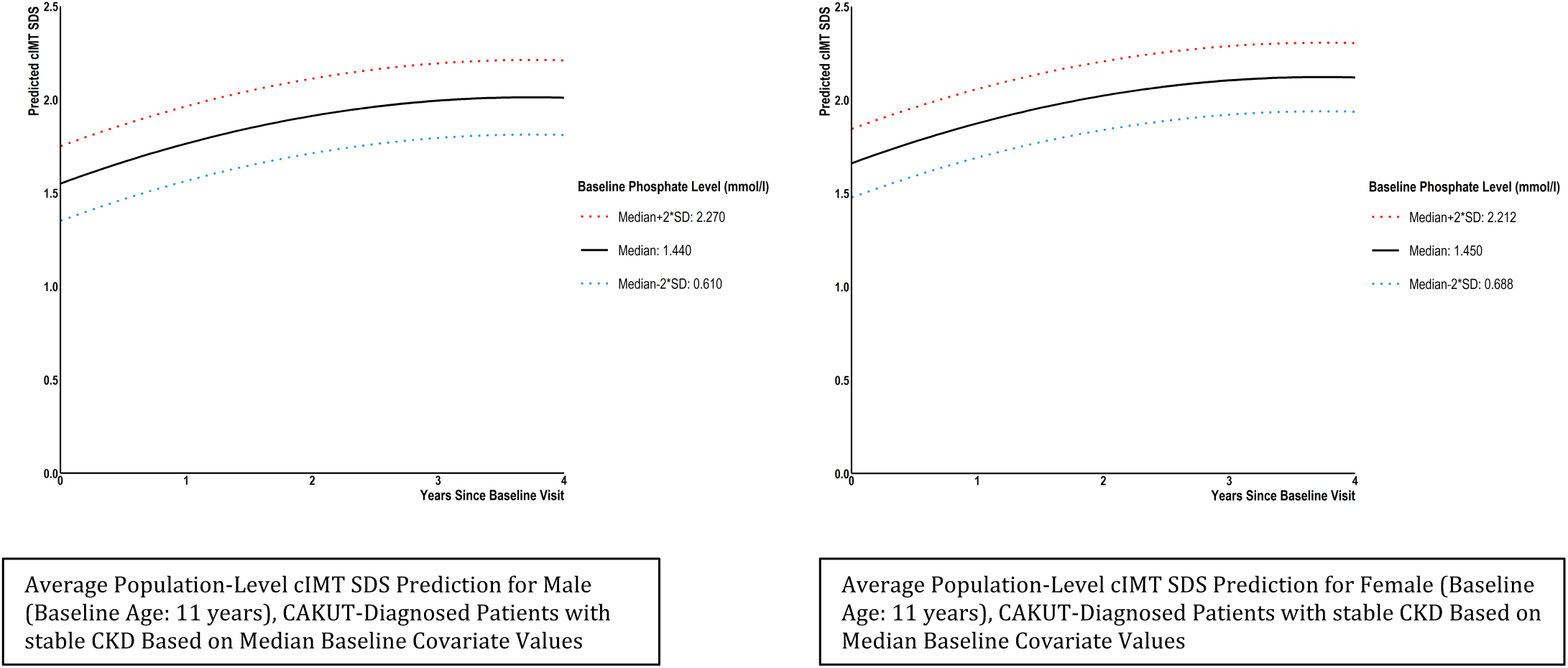
Average Population-Level cIMT SDS Prediction for Male and Female (Baseline Age: 11 years), CAKUT-Diagnosed Patients with stable Chronic Kidney Disease Based on Median Baseline Covariate Values

**Figure 4.**
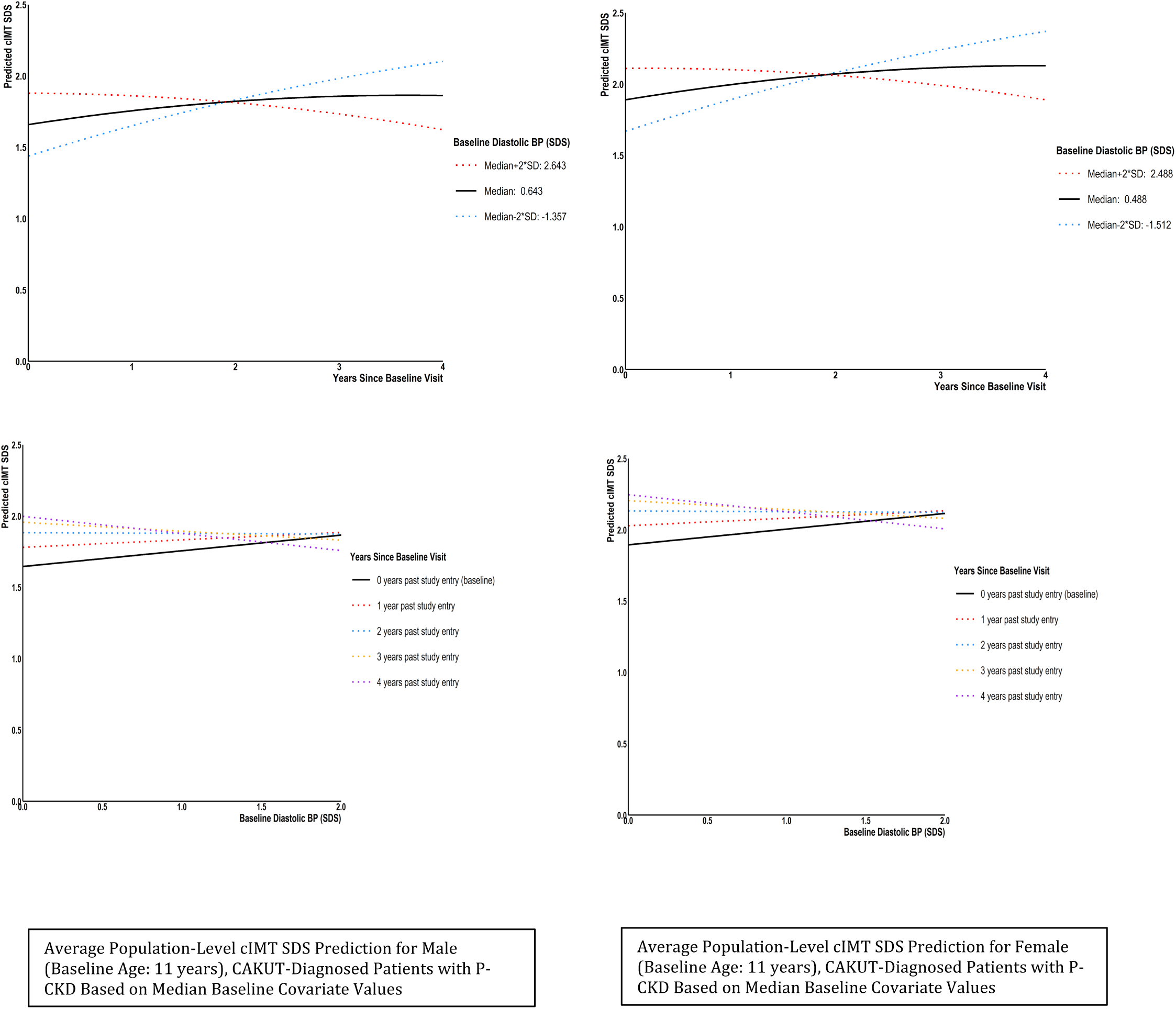
Average Population-Level cIMT SDS Prediction for Male and Female (Baseline Age: 11 years), CAKUT-Diagnosed Patients with Progressive Chronic Kidney Disease Patients Based on Median Baseline Covariate Values

**Table 4.** Pooled Population-Level (Fixed Effect) Mean Estimates of the Linear Mixed Model (LMM) with Modeled Time-Dependent Residual Variance Function for cIMT SDS for Patients with Stable Kidney Function.

| Fixed effect | $\beta$ | SE | CI95% | p |
| --- | --- | --- | --- | --- |
| Intercept | 12.980 | 0.3600 | 0.5915 2.0046 | .0003 |
| eGFR | 0.0078 | 0.0043 | -0.0007 0.0163 | .0733 |
| Log(albuminuria) | -0.0232 | 0.0267 | -0.0756 0.0292 | .3858 |
| Systolic blood pressure (SDS) | 0.0202 | 0.038 | -0.0544 0.0947 | .5955 |
| Diastolic blood pressure (SDS) | 0.0682 | 0.0449 | -0.0200 0.1564 | .1294 |
| **Phosphate** | **0.2408** | **0.1110** | **0.0228 0.4587** | **.0304** |
| **Time** | **0.2473** | **0.0525** | **0.1443 0.3503** | **<.0001** |
| **Time\*time** | **-0.0331** | **0.0080** | **-0.0488 -0.0174** | **<.0001** |
| Sex (female) | 0.1019 | 0.1154 | -0.1246 0.3284 | .3774 |
| Age at study entry | -0.0241 | 0.0160 | -0.0555 0.0073 | .1322 |
| Diagnosis (glomerulopathies) | -0.1701 | 0.2528 | -0.6662 0.3260 | .5012 |
| Diagnosis (post-AKI) | -0.1780 | 0.2718 | -0.7115 0.3554 | .5126 |
| Diagnosis (tubulointerstitial) | -0.1534 | 0.1928 | -0.5317 0.2250 | .4265 |
| Diagnosis (other) | 0.2563 | 0.3451 | -0.4209 0.9336 | .4578 |

Remark: Joint explained proportion of total cIMT SDS variance by included fixed effects
Time: Time in years since first visit. Time\*time: quadratic effect of time in the model.
Diastolic blood pressure\*time: interactive term between diastolic blood pressure (SDS) and time

**Table 5.** Pooled Population-Level (Fixed Effect) Mean Estimates of the Linear Mixed Model (LMM) with Modeled Time-Dependent Residual Variance Function for cIMT SDS for Patients with Progressive Kidney Failure.

| Fixed effect | $\beta$ | SE | CI95% | p |
| --- | --- | --- | --- | --- |
| intercept | 20.702 | 0.2782 | 1.5243 2.6160 | <.0001 |
| eGFR | -0.0026 | 0.0042 | -0.0108 0.0056 | .5366 |
| **Log(albuminuria)** | **0.0498** | **0.0264** | **-0.0020 0.1015** | **.0596** |
| Systolic blood pressure (SDS) | 0.0453 | 0.0318 | -0.0172 0.1077 | .1552 |
| **Diastolic blood pressure (SDS)** | **0.1101** | **0.0521** | **0.0078 0.2124** | **.0349** |
| **Height (sds)** | **0.1047** | **0.0325** | **0.0409 0.1685** | **.0013** |
| **Time** | **0.1495** | **0.0495** | **0.0524 0.2465** | **.0026** |
| **Time\*time** | **-0.0154** | **0.0076** | **-0.0304 -0.0004** | **.0442** |
| **Diastolic blood pressure (SDS)\*time** | **-0.0575** | **0.0155** | **-0.0878 -0.0272** | **.0002** |
| **Sex (female)** | **0.2983** | **0.0935** | **0.1148 0.4818** | **.0015** |
| **Age at study entry** | **-0.0543** | **0.0143** | **-0.0824 -0.0262** | **.0002** |
| Diagnosis (glomerulopathies) | -0.0995 | 0.1747 | -0.4423 0.2432 | .5689 |
| Diagnosis (post-AKI) | -0.1383 | 0.1934 | -0.5177 0.2412 | .4748 |
| Diagnosis (tubulointerstitial) | -0.2146 | 0.1246 | -0.4591 0.0299 | .0853 |
| Diagnosis (other) | -0.0817 | 0.2162 | -0.5058 0.3424 | .7055 |

Remark: Joint explained proportion of total cIMT SDS variance by included fixed effects
Time: Time in years since first visit. Time\*time: quadratic effect of time in the model.
Diastolic blood pressure\*time: interactive term between diastolic blood pressure (SDS) and time

In the linear mixed model, the effects of the identified cardiovascular risk factors with statistical significance on average for all patients jointly explained 10.9% 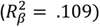 of the total cIMT SDS variance in the overall cohort. The remaining unexplained variance was decomposed by the linear mixed model into 47.0 % of inter-patient variability 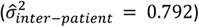, 11.2% of inter-center variability 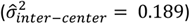, and into 41.8% of cIMT measurement error 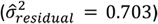 (see also supplementary Tables SVII – SIX).

Intra-patient correlations (IPC) indicated that baseline cIMT SDS measurements were moderately correlated with cIMT SDS measurements at 1 year of follow-up 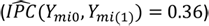. The estimated intra-patient correlation between cIMT SDS baseline and subsequent cIMT SDS measurements steadily decreased with follow-up time to an 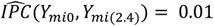 at 2.4 years of prospective follow-up. The dependence of measurements within patients validates LMM as a method of choice for modeling the longitudinal evolution of cIMT SDS within patients.

Building on the constructed linear mixed model, yearly progression rates of cIMT SDS (δ cIMT SDS) and blood pressure (δ diastolic BP and δ systolic BP) could be derived in order to evaluate whether dynamic changes of clinical factors predict progression of cIMT. These were calculated for the first 4.5 years of follow-up. There was a yearly relative change of cIMT SDS of 8.3 (−0.1, 20.6) %. The median relative change rate for systolic BP SDS was −5.2 (−11.4, 1.0) % and the relative diastolic BP SDS change per year was −6 (−11.3, −3.3) %. The association between dynamic blood pressure change (δ systolic and δ diastolic BP SDS) and cIMT SDS (δ cIMT SDS) rate were tested by linear regression analysis. This revealed a significant prediction of δ cIMT SDS by both δ systolic BP SDS (ß=041 ± 0.1, p<0.001, r^2^=0.03) and δ diastolic BP SDS (ß=1.56 ± 0.28, p<0.001, r^2^=0.06), indicating a dynamic association between blood pressure changes and cIMT changes over time (Figure 5).

**Figure 5.**
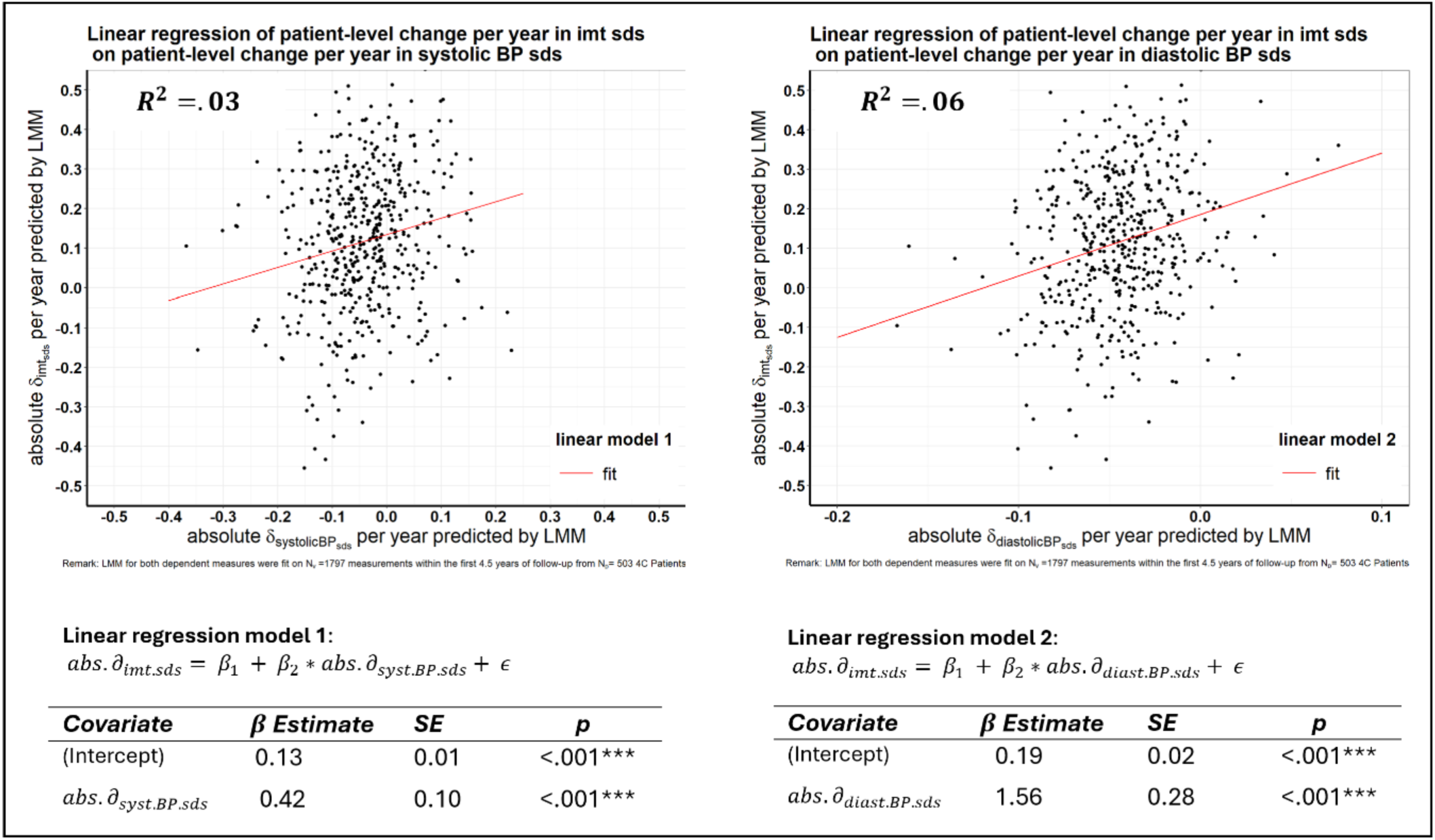
Progression of carotid intima media thickness (cIMT) depending on BP SDS dynamics over time: Test for Linear relationships between linear absolute change per year in cIMT SDS with linear absolute change per year in systolic and diastolic blood pressure SDS within first 4.5 years of follow-up. Abs.*δ*_imt.sds_= absolute change of cIMT SDS per year. Abs.*δ*_systolic.BP.sds_= absolute change of systolic BP SDS per year. Abs.*δ*_diast.BP.sds_= absolute diastolic BP SDS change per year. BP: blood pressure, SDS: standard deviation score

## Discussion

This is the first prospective study of the evolution of cIMT in a large cohort of children with CKD over the course of 8.5 years. We demonstrate a clear increase of cIMT standard deviation scores during the observation time, implying increasing cardiovascular risk over time in children with CKD when compared to their healthy peers. Associations of several established general and CKD-associated cardiovascular risk factors with cIMT SDS were observed, namely diastolic blood pressure, serum phosphate, serum albumin and albuminuria.

The rise in cIMT SDS was attenuated during extended observation time. However, start of dialysis or transplantation reduced patient numbers in the later visits, and this informative dropout may have contributed bias to the estimates of time effects. Also, there has been evidence that patients at a younger age have a higher impact of CKD on their vessels than later (see below). Our cohort might therefore mirror this crucial phase of growth, development and varying impact of detrimental effects according to age.

The **study design** and the distribution of patients across more than 50 centers required thorough planning and a vigorous joint effort of all participating centers to timely conduct synchronized visits and cardiovascular measurements. The enrolment of 700 patients with the rare condition of advanced pediatric CKD and the successful execution of the long-term study was made possible by the eminent clinical research experience and close collaborative ties of the ESCAPE Network centers. Besides pulse wave velocity and echocardiographic exams, cIMT measurement was chosen since it is an age-appropriate non-invasive measurement well suited to find and describe early alterations of the vascular system^17^. In particular, the repetitive performance of annual cardiovascular assessments by jointly trained regional investigators at their individually assigned study sites with portable devices was crucial to minimize observer-related variation of measurements in this multi-center, multi-year study. Hence, the 4C Study cohort constitutes a **unique dataset** in terms of underlying aligned methodology, cohort size, and realization.

cIMT SDS was clearly increased at the beginning of the study and there was further progression with time, even after statistically controlling for other associated risk factors. While several previous studies observed increased cIMT in both adult and pediatric CKD patients, the dynamics of cIMT in the course of disease is largely unknown due to the almost complete absence of longitudinal studies^18^. Our study covers a **time period of more than 8** years during a crucial phase of growth and development. The continuous increase in age-and sex adjusted cIMT measurements (i.e. cIMT-SDS) observed during the first 4 years slowed towards the end of the observation period, which was modeled by a positive linear and a negative quadratic effect in the LMM equation. This evolving progression over time shows that the structural transformation of large arteries in childhood-onset CKD is a dynamic process and implies that much of the damage occurs during the early phase of CKD and further aggravates before reaching ESKD. This is also supported by the CKiD study, which observed elevated cIMT in early CKD which was related to hypertension and dyslipidemia^19^. Similar findings have been made in adult CKD patients. Desbien et al. found that within two years of follow-up the largest increase of cIMT occurred in CKD patients with the lowest initial serum creatinine^20^.

Our longitudinal analysis identified **female sex and young age** as predictive factors for increased cIMT. While intriguing, the higher risk for increased cIMT at a younger age may reflect varying timespans of disease burden in younger patients predominantly being affected by CAKUT which will impact CKD starting from birth. Interestingly, the ARIC study in adults found increased hazard ratios for cardiovascular disease in patients younger than 65 when compared to older persons based on serum albumin-creatinine ratios. Here also it remained unclear why the overall risk declined with increasing age.^21^ The effect of sex was most prominent in the subgroup of patients with progressive kidney failure and corroborates parallel analyses of large-artery function in the 4C cohort. While younger women in general population seem to be better protected from cardiovascular disease^22^, in children with CKD of the 4C Study we consistently found higher levels and greater progression of elevated carotid-femoral pulse-wave velocity in girls compared to boys both prior to KRT and after initiation of dialysis or transplantation, especially so in those with faster eGFR decline and independently of the duration of CKD prior to ESKD.^5,7^ The ARIC study found a significantly increased risk for cardiovascular disease in female patients with increasing albumin-creatinine ratios.^21^ These findings add to the increasing understanding of sex differences in medicine^23^, particularly concerning CVD risk profiles and progression^24,25^. Once more, they advocate for sex-specific guidelines both in pediatric and adult medicine^22^.

Furthermore, in this longitudinal study we confirmed the pivotal role of **elevated blood pressure for early arterial damage**. Our findings are in accordance with those of the cross-sectional baseline analysis of the 4C study cohort^6^, in which we found systolic blood pressure, BMI SDS, female gender and serum phosphate as risk factors for higher cIMT SDS. An analysis of ambulatory blood pressure monitoring specifically identified night-time BP and isolated nocturnal hypertension as predictors of higher cIMT^26^. The longitudinal analysis of PWV also highlighted diastolic blood pressure as a relevant risk factor for increased vascular stiffness^7^, suggesting that diastolic BP may be more relevant for both functional and structural abnormalities of the cardiovascular system. This verifies and displays one of the mechanisms how CKD ultimately translates into CVD. While there is abundant evidence of the association of high blood pressure and cIMT from cross-sectional studies, the long-term impact of increased blood pressure on cIMT in CKD is largely unknown. One study in 70 children with type 1 diabetes mellitus distinguished blood pressure and BMI as a relevant factor for cIMT increase after 4 years^27^. Previous studies have used various means to describe the evolution of large artery morphology such as calculating a cardiovascular risk score at baseline and measuring cIMT after a certain observation period^28^ or by associating long-term BMI or blood pressure trajectories with cIMT in adulthood ^29,30^. Our study and analysis approach confirms their findings and adds further insight to the distinct impact of time and cardinal risk factors such as blood pressure, age and sex in CKD. Since other studies have described significant tracking of cIMT values from early youth well into adulthood, our findings are ever more consequential for the long-term health of pediatric CKD patients.

Interestingly, we found a significant association of diastolic BP with cIMT SDS in the subgroup of patients with progressive kidney failure but not in those with stable kidney function. This is in line with the analysis of Shroff et al., where children diastolic BP was a significant risk factor for increased cIMT in children approaching kidney replacement therapy^31^. We speculate that patients with stable kidney function are less fluid overloaded and will therefore be at lower risk to develop high cIMT as a consequence of increased diastolic BP. The interaction between time and diastolic blood pressure indicates that diastolic BP was less relevant for cIMT towards the end of the observation period.

Possibly, patients with fluid overload or high blood pressure were monitored more closely when kidney function declined while awareness for the effects of high BP increased continuously during the study period and treatment might have been intensified. This interpretation is in keeping with the described observation of intensifying antihypertensive therapy and decreasing systolic and diastolic BP during the study period.

**Reduced serum albumin** in the overall cohort, and albuminuria in the progressive CKD group, were significantly and time-independently associated with cIMT SDS. Both pro- and anti-calcification processes within the serum are altered in CKD^32,33^. Low serum albumin levels are even associated with a more rapid loss of GFR and have been linked to a higher incidence of cardiovascular events in CKD patients.^34,35,36^. They are a well-known risk factor for vascular calcifications and cardiovascular mortality in adult dialysis^37,38^, and predialysis patients^39^ and have been found significantly associated with increased cIMT in a study of children treated with peritoneal dialysis^40^. Moreover, low serum albumin is associated with a higher calcification propensity, a strong predictor of all-cause mortality in adults with predialysis CKD^41^. Recognized as a marker of inflammation and protein-energy wasting in CKD^42^, albumin may also have an active role in vascular remodeling. Recent in-vitro studies have shown that albumin protects vascular smooth muscle cells from toxic effects of calcium phosphate nanoparticles^43^.

Adding to the abundant literature about the deleterious effect of high **serum phosphate** in CKD on vessel morphology^18,44,45^, a recently published analysis from the CRIC study confirmed a positive association between serum phosphate levels and coronary artery calcification in a large population of adult CKD patients.^46^. Interestingly, we observed an impact of serum phosphate only in the group with stable stable CKD. In the group with progressive CKD, the importance of blood pressure and other factors may have outweighed the impact of phosphate.

While **kidney function** was not a significant predictor of cIMT in the total study cohort, we found different risk profiles for cardiovascular disease within our population depending on the stage and progression of CKD. This might indicate that concomitant factors of progressive CKD play a more important role for cardiovascular disease than eGFR decline.

The association of the dynamics of cIMT SDS and blood pressure is an important finding and may imply that changes of blood pressure over time in fact influence the progression of cIMT. In pediatric CKD patients, a small study in children with hypertension has shown that lowering of BP by antihypertensive treatment led to a decrease of cIMT after 1 year of follow-up^47^. While longitudinal data in larger groups in this regard is scarce in children, a large meta-analysis in adults by Willeit et al. has summarized over 119 controlled trials showing indeed that therapeutic intervention could reduce cIMT progression, leading ultimately to a reduction of cardiovascular disease^48^. While our findings are observational only, they add to hypothesis of a potential benefit of blood pressure control on target organ damage and therefore support the promotion of a controlled clinical trial to prevent cIMT progression by strict control of clinical risk factors such as BP.

The reasons for the decelerating increase of cIMT towards the end of the study are speculative. The almost decade-long study per se may have positively influenced the awareness for secondary complications of CKD in participating centers ultimately resulting in intensified treatment. The demonstrable effect of decreasing diastolic blood pressure in the cohort could possibly be explained by this effect, consecutively contributing to a slowing of the progression of cIMT SDS instead of the expected ‘natural’ continuous course of large artery vasculopathy.

### Limitations

This is the largest longitudinal study of cIMT and its progression in children with CKD to date. However, it is observational only and all assumptions and conclusions are based on correlational associations. As our analysis indicated a relevant measurement error, additional clinical effects might have been missed.

The unexplained variability in the regression model may stem from risk factors of clinical relevance at the individual patient level albeit not on the population level. Such inter-patient variation could for example derive from differential genetic susceptibility. This highlights the need for further research to explain the individual course of CVD progression, which could add to the advance of personalized medicine. And although environmental exposure to certain unmonitored modifying risk factors may have been missed, **important modifiable risk factors** such as blood pressure, serum albumin and albuminuria were among the identified predictive factors and, even more importantly, there seems to be a dynamic association between blood pressure modification and the progression of cIMT. This once again points to the benefits of a strict control of modifiable risk factors such as blood pressure. In this regard, an increasing awareness for secondary complications of CKD of the attending physicians over time and their modification by enhanced treatment may have had influence on the progression of cIMT on the population level.

Furthermore, the fact that cIMT SDS progressively increased over time should prompt further research for additive and interacting population-based risk factors and protective factors with clinical relevance for cIMT that could not be unraveled by the utilized LMM approach.

### Conclusions

In conclusion, this prospective long-term observational study of a large pediatric CKD cohort reveals an increased cIMT SDS at study entry which significantly progresses during the study follow-up, with an attenuation of the progression towards the end of the study. Several traditional cardiovascular risk factors could be identified as predictors for increased cIMT SDS, such low serum albumin, increased diastolic blood pressure and serum phosphate. Younger patients were significantly more at risk for increased cIMT SDS and there was a substantial sex difference especially for patients with progressive CKD with increased cIMT SDS in female patients. Importantly, there was a relationship between the yearly blood pressure change rate as a modifiable risk factor and cIMT SDS progression. Therefore, identification and treatment of modifiable cardiovascular risk factors should therefore be emphasized in all stages and ages of pediatric CKD. Further research should also address yet unidentified average population-level and patient-specific risk factors and the impact of treatment of modifiable risk factors such as blood pressure on individual progression of cIMT. Our findings highlight that all pediatric CKD patients, independent of CKD stage, should be screened for modifiable cardiovascular risk factors and treated accordingly, since this population is, and will be later in life, particularly prone to cardiovascular complications.

## Supporting information

Supplemental Material - Statistical Methods

## Data Availability

All data produced in the present study are available upon reasonable request to the authors

## Acknowledgements

We thank all children and adolescents and their parents for their participation in the study. We gratefully acknowledge the support of our colleagues and friends.

Support for the 4C Study was received from the ERA-EDTA Research Programme, the KfH Foundation for Preventive Medicine and the German Federal Ministry of Education and Research (01EO0802).

Several authors are members of the European Rare Kidney Disease Reference Network (ERKNet).

## Supplemental Material (see separate file)

Statistical Methods

Figure S1

Figure S2

Figure S3

Figure S4

Figure S5

Figure S6

References 12,13,15,16,49-53

## Non-standard Abbreviations and Acronyms

CVD: Cardiovascular Disease
CKD: Chronic Kidney Disease
4C: Study the Cardiovascular Comorbidity in Children with CKD study cIMT Carotid Intima-media Thickness
ESKD: End-stage Kidney Disease
PWV: Pulse Wave Velocity
KRT: Kidney Replacement Therapy
SDS: Standard Deviation Score
iPTH: intact Parathyroid Hormone
CRP: C-reactive Protein
eGFR: estimated Glomerular Filtration Rate
BP: Blood Pressure
WHO: World Health Organization
SD: Standard Deviation
LMM: Linear Mixed Model
P-CKD: Progressive Kidney Disease
S-CKD: Stable Chronic Kidney Disease
AKI: Acute Kidney Injury
IPC: Intra-patient Correlations
δ systolic BP SDS: yearly change rate of systolic BP SDS
δ diastolic BP SDS: yearly change rate of diastolic BP SDS

