## Supplemental Material - Statistical Methods for "Progression of Carotid Intima-Media Thickness in Children of the Cardiovascular Comorbidity in Children with Chronic Kidney Disease Study (4C Study) – Risk Factors and Impact of Blood Pressure Dynamics"

Anke Doyon[1], Jonas Hofstetter[1], Aysun Karabay Bayazit[2], Karolis Azukaitis[3], Ana Niemirska[4], Mahmut Civilibal[5], Ipek Kaplan Bulut[6], Ali Duzova[7], Berna Oguz[8], Bruno Ranchin[9], Rukshana Shroff[10], Yelda Bilginer[7], Salim Caliskan[11], Dusan Paripovic[12], Cengiz Candan[13], Alev Yilmaz[14], Jerome Harambat[15], Z. Birsin Özçakar[16], Francesca Lugani[17], Harika Alpay[18], Sibylle Tschumi[19], Ebru Yilmaz[20], Dorota Drozdz[21], Yilmaz Tabel[22], Gül Özcelik[23], Alberto Caldas Afonso[24], Onder Yavascan[25], Anette Melk[26], Uwe Querfeld[27], Franz Schaefer[1], for the 4C Study Consortium

[1] Pediatric Nephrology Division, Center for Pediatrics and Adolescent Medicine, Heidelberg, Germany
[2] Department of Pediatric Nephrology, Çukurova University, Faculty of Medicine, Adana, Turkey
[3] Clinic of Pediatrics, Institute of Clinical Medicine, Faculty of Medicine, Vilnius University, Vilnius, Lithuania
[4] Department of Pediatric Nephrology, The Children's Memorial Health Institute, Warsaw, Poland
[5] Department of Pediatric Nephrology, Haseki Educational and Research Hospital, Istanbul, Turkey
[6] Pediatric Nephrology Division, Department of Pediatrics, Ege University Medical Faculty, Izmir, Turkey
[7] Division of Pediatric Nephrology, Dpt. Of Pediatrics, Hacettepe University Faculty of Medicine, Ankara, Turkey
[8] Department of Radiology, Hacettepe University Faculty of Medicine, Ankara
[9] Pediatric Nephrology Division, Hospices Civils de Lyon, Hôpital Femme-Mère-Enfant, Centre de référence de maladies rénales rares, Université de Lyon, Bron, France
[10] Renal Unit, Great Ormond Street Hospital for Children NHS Foundation Trust, London, UK
[11] Istanbul University Cerrahpasa, Cerrahpasa Medical Faculty, Pediatric Nephrology, Istanbul
[12] Nephrology Department, University Children's Hospital and School of Medicine, University of Belgrade, Serbia.
[13] Division of Pediatric Nephrology, Istanbul Medeniyet University, Faculty of Medicine, Istanbul, Turkey
[14] Pediatric Nephrology, Istanbul Medical Faculty, Istanbul, Turkey
[15] Pediatric Nephrology Unit, Department of Pediatrics, Bordeaux University Hospital, France
[16] Division of Pediatric Nephrology, Department of Pediatrics, Ankara University Medical School, Ankara, Turkey
[17] Pediatric Nephrology, Istituto Giannina Gaslini, Genoa, Italy
[18] Pediatric Nephrology, Marmara University Medical Faculty, Istanbul, Turkey
[19] Children's Hospital, Inselspital, Bern, Switzerland
[20] Sanliurfa Children's Hospital, Sanliurfa, Turkey
[21] Department of Pediatric Nephrology and Hypertension, Jagiellonian University Medical College, Krakow, Poland
[22] Pediatric Nephrology, Uludag University, Bursa, Turey
[23] Department of Pediatrics, Division of Pediatric Nephrology, SBÜ.Şişli Hamidiye Etfal Training and Research Hospital, İstanbul, Turkey
[24] Pediatric Nephrology, Hospital Sao Joao, Porto, Portugal
[25] Department of Pediatrics, Division of Pediatric Nephrology, Medipol University, Faculty of Medicine, 34214 Istanbul, Turkey.
[26] Department of Pediatric Kidney, Liver and Metabolic Diseases, Hannover Medical School, Hannover, Germany
[27] Department of Pediatrics, Division of Gastroenterology, Nephrology and Metabolic Diseases, Charité-Universitätsmedizin Berlin, 13353 Berlin, Germany.

Running title: cIMT Progression in Children with CKD (4C Study)

Content:

Statistical Methods

Figure S1-S6

### Data Exploration

cIMT SDS across patients and visits followed a normal distribution (Figure S1A). Linear Locally Estimated Scatterplot Smoothing (LOESS) revealed an average quadratic, concave-down evolution of cIMT SDS with a more pronounced increase within the first 4 years of follow-up for patients with S-CKD compared to patients with P-CKD (Figure S1B). Patients dropped out disproportionally often due to start of kidney replacement therapy during the first 4 years of follow-up (Figure S1C). The last visit of patients with progressive CKD occurred on average 244 days (SD = 228 days) prior to RRT onset with a mean eGFR = 14.616 (SD = 6.153) at their last included visits. The last included visit of patients with stable CKD within the first 4 years of follow-up showed a mean eGFR = 30.494 (SD = 12.839) corresponding to CKD stage 4.

A quadratic decline in cIMT SDS LOESS trajectories after 4 years of follow-up (Figure S1B) was equally present for P-CKD and S-CKD patients and coincided with decreasing numbers of patients who contributed data at longer follow-up times (Figure S1C). Patients with total 4C participation durations > 5 years showed decreased cIMT SDS at their last visit compared to the last visits of patients with total 4C participation durations < 5 years (Figure S1D). The correlation of longer follow-up times with  lower cIMT SDS at last visits may be indicative for an informative drop-out mechanism (Missing Not At Random, MNAR) resulting in missing data[49] increased cIMT SDS are predictive for patient drop-out. If higher values on cIMT SDS are more likely to be missing, LMM fixed effect estimates are biased downwards because they are based on the lower values that are more likely to be observed. The impact of a MNAR underestimation bias was assumed to be small as informative dropout appeared to be a temporally confined issue of follow-up durations > 5 years where relatively few data remained to impose bias on fixed effect estimates (Figure S1D).

The empirical variance of cIMT SDS was dependent on patients´ time of follow-up with greatest observed cIMT SDS variance at baseline and continuously decreasing variance for longer follow-up durations (see Figure S1B). The greatest decrease in cIMT SDS variance occurred between patients´ 1$^{st}$ (baseline) visit and patients´ first annual follow-up visit (see Figure S1B & Table 1). Individual longitudinal patient profiles showed non-linear (quadratic) time trends (Figure S1E).

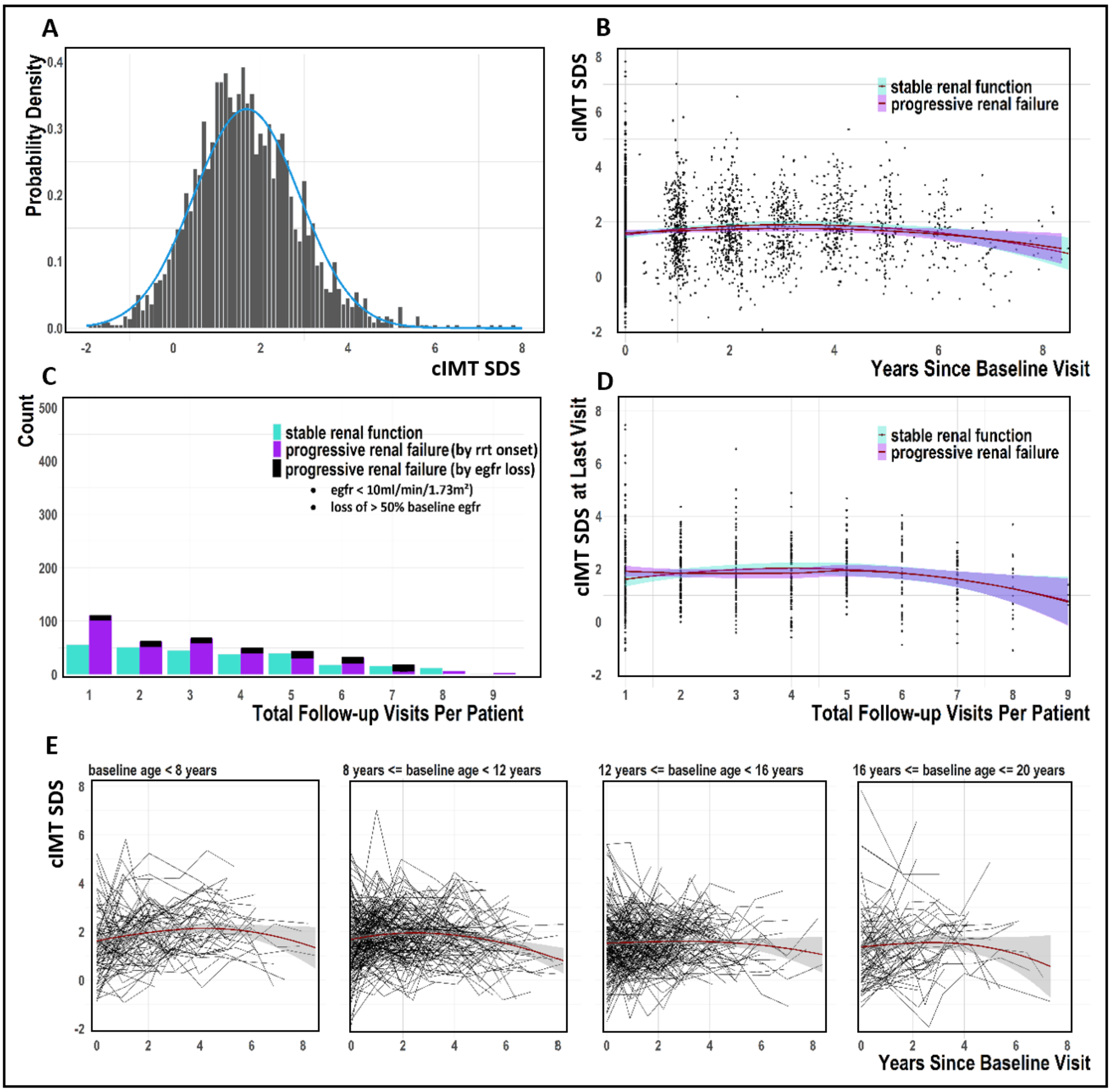


**Figure S1**: Visual exploration of cIMT SDS. **A**: Frequency distribution of cIMT SDS with overlayed normal density function (blue line). **B**: Scatterplot of cIMT SDS over time (in years) since baseline visit with linear LOESS trends separately for patients with PRF and SRF (span = 2, shading indicates +/- 1 SE). **C**: Count of patients with specific total number of visits (visit 1 = baseline visit) separately for patients with progressive CKD (differentiated by categories for patient inclusion) and stable CKD. **D**: cIMT SDS at patients´ last visit for patients´ with different total numbers of visits. Linear LOESS trend (span = 2) of this relationship is plotted separately for patients with PRF and SRF (shading indicates +/- 1 SE). **E**: cIMT SDS over time (in years) since baseline visit for single patients (black lines) and different groups of starting age at baseline (separate panels) with linear average LOESS trend (span = 2, shading indicates +/- 1 SE).

### Multiple Imputation of Missing Values

The percentage of missing values was below 10% for most explanatory variables and there was only 1 single missing cIMT measurement (Figure S2A). Missing values of all explanatory covariates and of cIMT SDS were imputed to generate twenty complete (imputed) datasets (m = 20). The lmer.ml function (from the mice[50] and miceadds[51] R packages) with *predictive mean matching (PMM)* method specification was applied to the preprocessed 4C dataset to conduct multiple imputation in R Studio v. 4.2.2[16,15] . To generate each imputed dataset, lmer.ml fitted a linear mixed model for each variable with missingness, by including all remaining covariates in the fixed effect predictor set. Only covariate main effects were estimated. The LMM was specified with center-level random intercepts and patient-level random intercept and random slopes. Based on the LMM predicted patient-level profiles, lmer.ml applied the PMM method. PMM substitutes each missing covariate value for a particular visit and patient with a non-missing and randomly drawn observed value (donor value) in close distance to the prediction for the missing value. The utilized PMM default hyperparameter value k = 5 limited the candidate pool for the randomly drawn donor value to the five closest values available across all visits and patients. PMM ensures that the resulting imputed values are within the range of the observed non-missing values and no outliers are generated[52] . Figure S2B exemplifies that PMM (k=2) already reliably identifies donor values (red dots) that maintain the overall shape of a patient-level covariate trajectory. The fixed effect estimates and estimates of random effect covariance parameters obtained by the final inferential LMM per patient group on each of the 20 imputed datasets were pooled according to Rubin's Rule[13] and are reported in the result section of this paper.

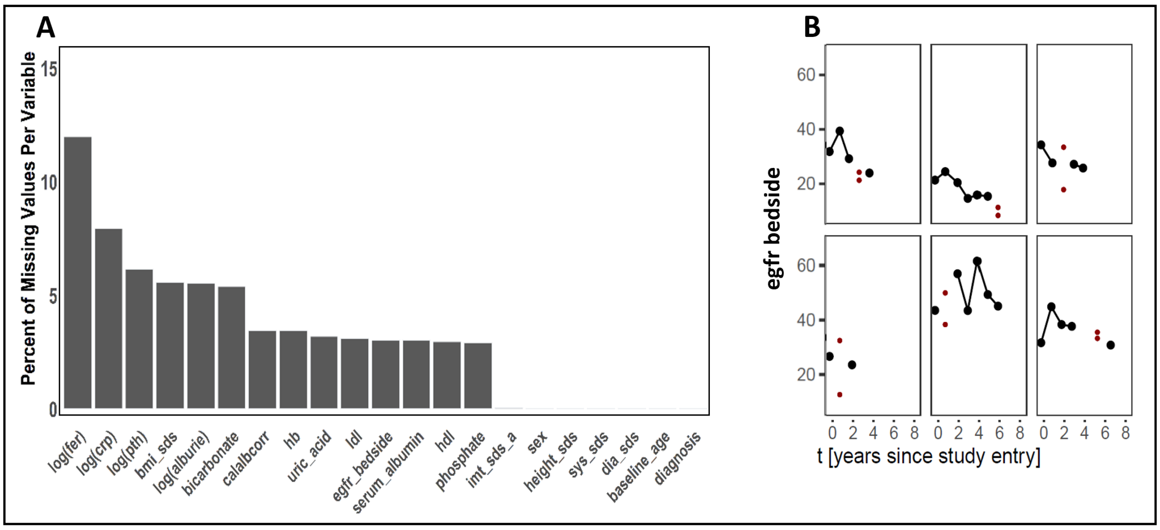


**Figure S2**: **A**: Percent of missing values for cIMT SDS and per explanatory covariate of interest. **B**: Exemplary results of PMM-based (k=2) multiple imputation obtained from one (m=1) imputed 4C analysis dataset. Each of the six subpanels displays observed data (black dots) for one 4C patient and the pool of donor values determined by PMM (2 red dots) out of which one value is selected for missing value substitution.

### An Application-Specific Linear Mixed Model Formulation

Separate LMM were specified for longitudinal modeling of cIMT SDS in all patients, in patients with progressive CKD (P-CKD), and in patients with stable CKD (S-CKD).

All three LMM were first constructed with a complex mean structure that included all single explanatory covariates of interest in covariate matrix $\boldsymbol{X}$ for the estimation of time-invariant, and first-degree time-variant fixed effects (i.e. average population-level effects). LMM fixed effects can be interpreted as characterizing the longitudinal relationship between explanatory covariates (CVD risk factors) and cIMT SDS for the *"average"* patient, given a specific configuration of explanatory covariate values. In addition to continuous time (i.e. time in years of follow-up after baseline), and time, explanatory covariates in the complex mean structure included: eGFR, serum albumin, HDL, LDL, phosphate, albumin-corrected calcium, serum bicarbonate, uric acid, log(ferritin), log(CRP), log(iPTH), log(albuminuria), hemoglobin, BMI SDS, height SDS, systolic blood pressure SDS, diastolic blood pressure SDS, baseline age, diagnosis, sex. Right-skewed covariates were added as log-transformed covariates. Each explanatory covariate was added as a single covariate to the three initial LMM formulations and in interaction with the continuous covariate time to assess whether the association between a covariate and cIMT SDS varied over time. Multicollinearity between all covariates and interaction terms was examined which resulted in the exclusion of the following four interaction terms due to almost perfect multicollinearity based on Pearson correlation coefficients ($r > .95$), and Variance Inflation Factors ($VIF > 20$): albumin-corrected calcium * time, serum albumin * time, hemoglobin * time, serum bicarbonate * time. The initial, preliminary complex mean structure was then kept unchanged during Restricted Maximum Likelihood (REML) estimation of an LMM for each patient group while systematically varying the specified random effect covariance structure of each model (Table S1-S3). The selection of an appropriate and best-fitting random effect covariance structure per LMM, firstly, aimed at explicitly modeling how individual patients and centers deviate from estimated average longitudinal population-level effects and, secondly, took into account longitudinal covariance (dependence) of cIMT SDS measurements within patients, and between patients from the same treating center. The set of probed random effect covariance structures was informed by the general knowledge about the 4C registry data (i.e. allowing centers´ individual mean cIMT SDS at baseline to deviate from the overall population-level mean seemed reasonable) and by results of the data exploration (quadratic patient-level random effects were supported by visualizations of patient-specific longitudinal profiles; see supplemental method sections). The decision about the best LMM random effect structure was then guided by Likelihood Ratio Tests (LRT) during model comparison and by the Akaike Information Criterion (AIC) which penalizes improved LMM fit by a model´s parameter complexity[12] . Table SI-SIII show that a random effect covariance structure specification with center-level random intercepts, and patient-level random intercepts, patient-level linear random slopes, and patient-level quadratic random slopes resulted in the best model fit as indicated by AIC and LRT for LMM of cIMT SDS in all three patient groups (see LMM Model #7 in Tables SI-SIII). Figure 2.1 exemplifies that especially patients´ whose longitudinally observed cIMT SDS profile (black dots) diverged more distinctly from the estimated longitudinal average population-level trend (red line) can be predicted more accurately by LMM if, additionally to a standard inclusion of patient-level random intercepts and center-level random intercepts (see green lines in Figure 2.1 A-D), patient-level random linear slopes (see green lines in Figure 3 E-H), or, patient-level random linear slopes and random quadratic slopes (see green lines in Figure 2.1 I-L) are modeled by the LMM. The LMM Model #7 random effect covariance structure was therefore selected for the longitudinal LMM of cIMT SDS in all three patient groups (see Tables SI-SIII).

The standard LMM formulation assumes constant residual variance across all values of included explanatory covariates (homoscedasticity assumption). Fixed effect estimation precision decreases if the homoscedasticity assumption is violated. Data exploration showed evidence of exponentially decreasing cIMT SDS variance for increasing time of follow-up. It was considered that the observed time-dependent variance function may not be exclusively explainable by longitudinally converging patient-specific cIMT SDS profiles (patient-level random slopes) but may (additionally) be evoked by unequal amounts of measurement error (residual variance) over time. Thus, it was tested if explicit modelling of non-constant residual variance (measurement error) over time improved the LMM fit. Table I-III show that modelling non-constant residual variance over time improved model fit of LMM #7 in all patient groups. The REML-estimated negative value of the exponential parameter $\hat{\delta}$ in Tables I-III translates into an exponential factor $0 < e^{\hat{\delta} t_{ij}} < 1$ which is multiplied by the estimated residual variance $\widehat{\sigma^2}_{res}$ at follow-up times $t_{ij}$ greater than 0 (baseline). Hence, as expected, the residual variance estimate $\widehat{\sigma^2}_{res}$ was estimated to become smaller for increasing follow-up time by multiplication with $e^{\hat{\delta} t_{ij}}$.

After selecting a random effect covariance structure and confirming the need for modeling non-constant residual variance over time, the complex mean structure of LMM #7 was reduced by p-value based variable backwards elimination (covariate inclusion threshold; $p \leq .05$) to minimize the risk of fixed effect model overfitting to spurious correlations and random noise. Covariates of primary interest and covariates for effect adjustment were defined and never removed during variable backwards elimination. The list of single covariates of primary interest included: eGFR, log(albuminuria), systolic blood pressure SDS, diastolic blood pressure SDS. The list of single covariates always kept for fixed effect adjustment included: sex, age at study entry, diagnosis. All other covariates and interaction terms in the LMM #7 were candidates for removal during variable backwards elimination. The estimated average cIMT SDS population-level trend (red line) estimated by the final LMM with reduced fixed effect mean-structure is visualized in Figure 2.2 A-D for the visit-specific covariate values of four example patients.

$$\boldsymbol{Y}_{mij} = \underbrace{\beta_0 + c_{0m} + b_{0i}}_{\substack{\text{intercept}\\ \text{of patient } i\\ \text{in center } m}} + \underbrace{(\beta_1 + b_{1i})}_{\substack{\text{linear slope}\\ \text{of patient } i\\ \text{from } ti(j\text{-}1) \text{ to } t(j)}} t_{ij} + \underbrace{(\beta_2 + b_{2i})}_{\substack{\text{quadratic slope}\\ \text{of patient } i\\ \text{from } ti(j\text{-}1) \text{ to } ti(j)}} t^2{}_{ij} + \underbrace{\widetilde{\boldsymbol{X}}'_{ij}\widetilde{\boldsymbol{\beta}}}_{\substack{\text{remaining}\\ \text{fixed effects}\\ \text{(time invariant)}}} + \underbrace{\breve{\boldsymbol{X}}_{ij} t_{ij} \breve{\boldsymbol{\beta}}}_{\substack{\text{remaining}\\ \text{fixed effects}\\ \text{(time variant)}}} + \epsilon_{ij}$$

**where**:

$$b_i = [b_{0i}, b_{1i}, b_{2i}] \sim N(0, \boldsymbol{D}),\; c_m = [c_{0m}] \sim N(0, \boldsymbol{G}),\; \epsilon_{ij} \sim N(0, \boldsymbol{\Sigma}),$$

$$\boldsymbol{D} = \begin{bmatrix} var(b_0) & cov(b_0, b_1) & cov(b_0, b_2) \\ cov(b_0, b_1) & var(b_1) & cov(b_1, b_2) \\ cov(b_0, b_2) & cov(b_1, b_2) & var(b_2) \end{bmatrix} = \begin{bmatrix} d_{oo} & d_{01} & d_{02} \\ d_{01} & d_{11} & d_{12} \\ d_{02} & d_{12} & d_{22} \end{bmatrix},$$

$$\boldsymbol{G} = var(c_0) = g_{oo}\,\text{,}$$

$$\boldsymbol{\Sigma} = \boldsymbol{I}\,\sigma^2{}_{res} * e^{\delta t_{ij}} = \begin{bmatrix} \sigma^2{}_{res} * 1 & 0 & \cdots & 0 \\ 0 & \sigma^2{}_{res} * e^{\delta t_{i(j=2)}} & \ddots & 0 \\ \vdots & \ddots & \ddots & 0 \\ 0 & 0 & 0 & \sigma^2{}_{res} * e^{\delta t_{i(j=J)}} \end{bmatrix}.$$

*(equation I)*

The final longitudinal LMM per patient group is described by model equation I. In LMM equation I, the average population-level cIMT SDS intercept at baseline $\beta_0$, the linear average population-level effect $\beta_1$ of time on cIMT SDS, as well as the explanatory time covariate $t_{ij}$ are removed from the original regressor matrix $\boldsymbol{X}$ and from the original coefficient vector $\boldsymbol{\beta}$**,** respectively, to better illustrate the LMM formulation. Explanatory covariates with modeled time-invariant (stable) linear relationships with cIMT SDS of patient *i* from center *m* at individual follow-up time *j* ($\boldsymbol{Y}_{\boldsymbol{mij}}$) are contained in matrix $\widetilde{\boldsymbol{X}}$ to estimate the corresponding population-level average effects in vector $\widetilde{\boldsymbol{\beta}}$ between any times $t_{i(j-1)}$ and $t_{i(j)}$. Time-interacting explanatory covariates are included in matrix $\breve{\boldsymbol{X}}$ to estimate the linear rate of change $\breve{\boldsymbol{\beta}}$ in the covariates´ average linear relationship with cIMT SDS ($Y_{mij}$) between any times $t_{i(j-1)}$ and $t_{i(j)}$. The LMM in equation I models center *m´s* deviation from an estimated average longitudinal population-level trajectory, and patient *i´*s deviation from the modeled trajectory of his/her treating center *m*. Allowing for center-level random intercepts $c_{0m}$, and for patient-level random intercepts $b_{0i}$, patient-level random linear slopes $b_{1i}$, and patient-level quadratic slopes $b_{2i}$ requires the estimation of random effect variance parameter $g_{00}$ in the $\boldsymbol{G}$ matrix, and of 6 random effect (co)variance component parameters in the $\boldsymbol{D}$ matrix (see

$d_{00}, d_{11}, d_{22}, d_{01}, d_{02}, d_{12}$ in equation I). The estimated covariance components in ***G*** and ***D*** specify the predictive distribution for the center-level random effects $c_{0m}$, and for all patient-level random effects $b_{ki}$, respectively.

The final LMM per patient group (all patients, patients with P-CKD, patients with S-CKD) was fitted to 20 imputed datasets and the estimated LMM parameters were pooled according to Rubin's Rule[13] . The pooled fixed effect estimates, the pooled center-level and patient-level random effect covariance parameters, the derived cIMT SDS variability contributions at each hierarchical level of the model structure (average population-level, center-level, patient-level), and the derived *intra-patient-level* correlations for specific time intervals of follow-up are reported in the result section of this paper for the final LMM per patient group. The lme function from the nlme package[9] was utilized in R Studio vs 4.2.2[16,15] for longitudinal linear mixed modeling.

### Model Diagnostics

A plot per patient group´s final fitted LMM of the standardized marginal residuals against time of follow-up showed that modeling non-constant residual variance over time remedied residual heteroscedasticity during fixed effect estimation (Figure S3 D-E). Overall, the fitted average population-level longitudinal trends seemed to be free of systematic biases except for a small group of patients at follow-up durations > 6 years whose cIMT SDS were overestimated (Figure S3 D-E). Standardized marginal residuals appeared to be normally distributed across the entire range of average population-level fitted cIMT SDS suggesting that the LMM did not miss any apparent non-linear fixed-effect relationships between covariates and cIMT SDS in the data (Figure S3 A-C). Measurement errors within patients were not correlated between follow-up visits as shown by flat LOESS trend lines in semivariograms constructed for each fitted final LMM per patient group (Figure S3 G-I).

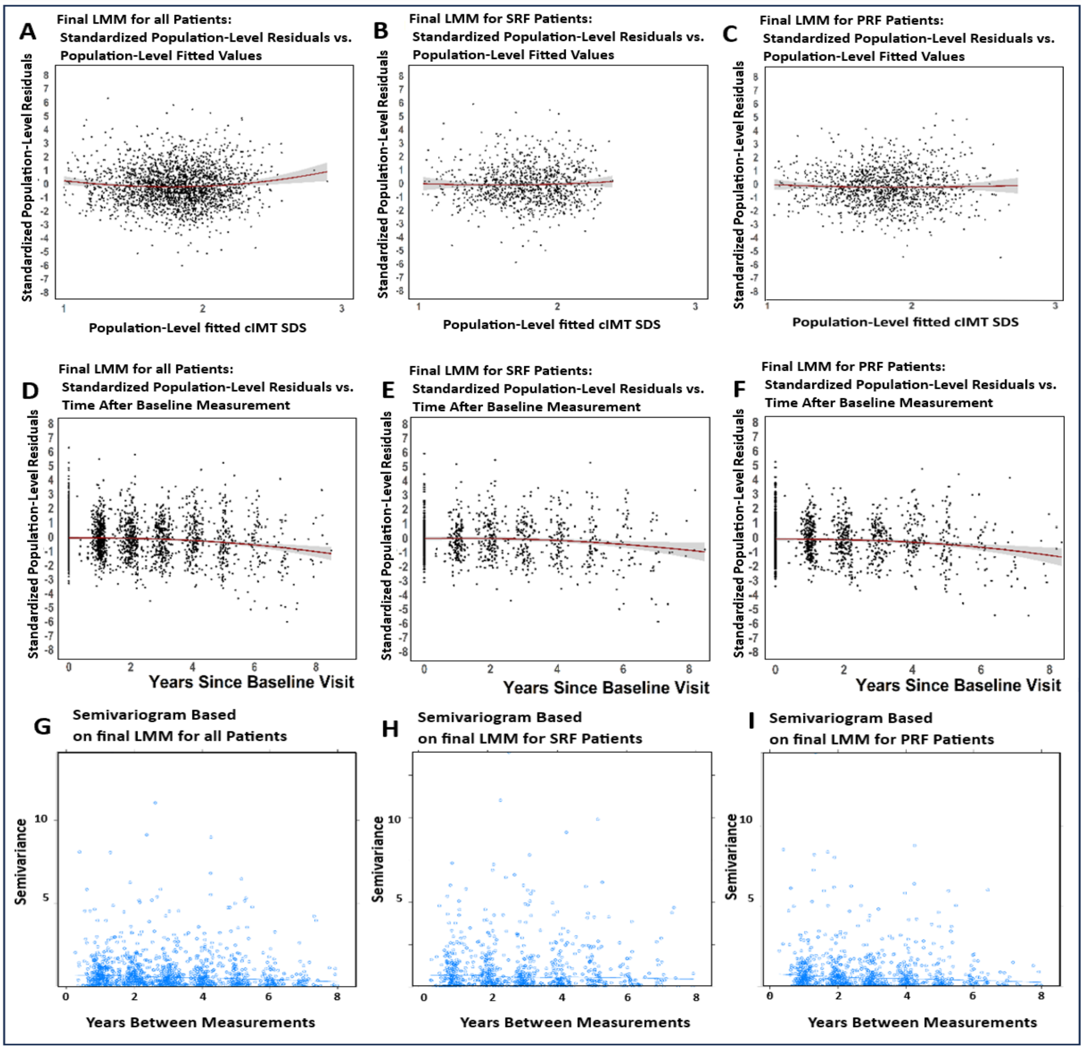


**Figure S3**: Model diagnostics for LMM per patient group. A-C: Plots of standardized marginal residuals against time of follow-up; D-F: Plots of standardized marginal residuals against marginal fitted cIMT SDS; G-I: Semivariograms.

### Derivations from the longitudinal LMM equation (equation I)

#### Decomposition of the estimated total cIMT SDS variance $\hat{\boldsymbol{\sigma}}^2_{\boldsymbol{j,total}}$

The decomposition of the estimated total cIMT SDS variance $\hat{\sigma}^2_{j,total}$ at time *j* around the average population-level mean $\boldsymbol{X'\hat{\beta}}$ into inter-center variability ($\hat{\sigma}^2_{inter-center}$), inter-patient variability ($\hat{\sigma}^2_{j,inter-patient}$), and time-dependent measurement error within patients ($\hat{\sigma}^2_{j,residual} = \sigma^2{}_{res} * e^{\delta t_{ij}}$) in the final LMM is represented by equation II.

$$
\begin{aligned}
\widehat{\mathrm{Var}}(Y_{mij}) &= \hat{\sigma}^2_{inter-center} + \hat{\sigma}^2_{j,inter-patient} + \hat{\sigma}^2_{j,residual} = \hat{\sigma}^2_{j,total} \\
&= \mathrm{Var}(\boldsymbol{X'\hat{\beta}} + \widehat{c_{0m}} + \widehat{b_{0\iota}} + \widehat{b_{1\iota}}t_{ij} + \widehat{b_{2\iota}}t^2{}_{ij} + \widehat{\epsilon_{\iota j}}) \\
&= \mathrm{Var}(\widehat{c_{0m}}) + \mathrm{Var}(\widehat{b_{0\iota}} + \widehat{b_{1\iota}}t_{ij} + \widehat{b_{2\iota}}t^2{}_{ij}) + \mathrm{Var}(\widehat{\epsilon_{\iota j}}) \\
&= \mathrm{Var}(\widehat{c_{0m}}) + \\
&\quad \mathrm{Var}(\widehat{b_{0\iota}}) + t^2{}_{ij}\mathrm{Var}(\widehat{b_{1\iota}}) + t^4_{ij}\mathrm{Var}(\widehat{b_{2\iota}}) + \\
&\qquad 2\mathrm{Cov}(\widehat{b_{0\iota}}, \widehat{b_{1\iota}}t_{ij}) + 2\mathrm{Cov}(\widehat{b_{0\iota}}, \widehat{b_{2\iota}}t^2{}_{ij}) + 2\mathrm{Cov}(\widehat{b_{1\iota}}t_{ij}, \widehat{b_{2\iota}}t^2{}_{ij}) + \\
&\quad \mathrm{Var}(\widehat{\epsilon_{\iota j}}) \\
&= g_{00} + \hat{d}_{00} + \hat{d}_{11}t^2{}_{ij} + \hat{d}_{22}t^4_{ij} + 2\hat{d}_{01}t_{ij} + 2\hat{d}_{02}t^2{}_{ij} + 2\hat{d}_{12}t^3_{ij} + \hat{\sigma}^2{}_{res} * e^{\hat{\delta}t_{ij}}.
\end{aligned}
$$

(*equation II*)

#### Assessment of the proportion of explained total cIMT SDS variance by the average population-level effects

To assess the proportion of explained total cIMT SDS variance by the average population-level effects , the standardized generalized variance $R^2_\beta$ metric (see equation III) proposed by Edwards et al. (2008)[53] was calculated for the final fitted LMM per patient group. The $R^2_\beta$ metric compared the final fitted LMM per patient group with the null model fit per patient group i.e. an LMM with identical random effect and residual covariance structures but intercept-only fixed effect mean structure.

$$
R^2_\beta = \frac{SSE_\beta(\boldsymbol{Y}_{mij} = \boldsymbol{1}\beta_0 + \boldsymbol{Z}_{ij}\boldsymbol{D}_{ij} + \boldsymbol{1}\boldsymbol{G}_i + \varepsilon_{ij}) \; - \; SSE_\beta(\boldsymbol{Y}_{mij} = \boldsymbol{X}_{ij}\boldsymbol{\beta} + \boldsymbol{Z}_{ij}\boldsymbol{D}_{ij} + \boldsymbol{1}G_i + \varepsilon_{ij})}{SSE_\beta(\boldsymbol{Y}_{mij} = \boldsymbol{1}\beta_0 + \boldsymbol{Z}_{ij}D_{ij} + \boldsymbol{1}\boldsymbol{G}_i + \varepsilon_{ij})}.
$$

(*equation III*)

### Intra-patient correlation (IPC)

The estimated *intra-patient correlation (IPC)* between any two cIMT SDS measurements of the same patient *i* at follow-up time $t_{i(j)}$ and follow-up time $t_{i(j+k;\, k\neq 0)}$ is expected to decrease with increasing time *k* between the two measurements and can be calculated based on the estimated LMM parameters as shown in equation IV.

$$\widehat{IPC}(Y_{mij}, Y_{mi(j+k;\, k\neq 0)}) = \frac{\widehat{Cov}(Y_{mij}\,, Y_{mi(j+k;k\neq 0)})}{\widehat{SD}(Y_{mij}) * \widehat{SD}(Y_{mi(j+k;k\neq 0)})}$$

$$= \frac{\hat{g}_{00} + \hat{d}_{00} + \hat{d}_{01}(t_{ij}+t_{i(j+k)}) + \hat{d}_{11}t_{ij}t_{i(j+k)} + \hat{d}_{02}({t^2}_{ij}+{t^2}_{i(j+k)}) + \hat{d}_{12}({t^2}_{ij}t_{i(j+k)}+t_{ij}{t^2}_{i(j+k)}) + \hat{d}_{22}{t^2}_{ij}{t^2}_{i(j+k)}}{\sqrt{\widehat{Var}(Y_{mij})} * \sqrt{\widehat{Var}(Y_{mi(j+k)})}}.$$

(*equation IV*)

### Post-fit correlations of absolute linear patient-level change per year in imt sds with absolute linear patient-level change per year in blood pressure sds within the first 4.5 years of follow-up

The analysis was conducted to investigate if there is a statistically significant relationship between estimated linear absolute rate of change in cIMT SDS with estimated linear absolute rate of change in blood pressure SDS within the first 4.5 years of follow-up. Patient-level absolute change per year is derived from the longitudinal linear mixed-effects model (LMM with REML estimation) constructed above and estimated for each of three dependent measures (cIMT SDS, systolic and diastolic BP SDS). Subsequently, two linear regression models are performed to test the correlation between the linear absolute change per year in cIMT SDS and either diastolic or systolic BP SDS.
Absolute change per year for patient *i* in systolic BP sds and in diastolic BP sds is obtained as (abs. $\delta$ .systolic.sds$_i$ = $\beta_1$ + b1$_i$ | abs. $\delta$ .diastolic.sds$_i$ = $\beta_1$ + b1$_i$) from estimated parameters of the corresponding LMM fitted to data from patients with stable renal function and progressive renal failure (see equation II).
Absolute change per year for patient *i* in imt sds is derived as (abs. $\delta$ .imt.sds$_i$ = (last.imt.sds$_i$ – first.imt.sds$_i$)/last.time$_i$) from predicted values of an LMM elaborated for testing risk factor relationships with imt sds in patients with stable renal function and progressive renal failure (see equation I in Appendix).

Predicted outcome and slope estimate values visualized in Figures S4-S6 are average-aggregated values based on repeated LMM fits to each of 20 imputed datasets.

**Patient-level absolute change per year in cIMT SDS**

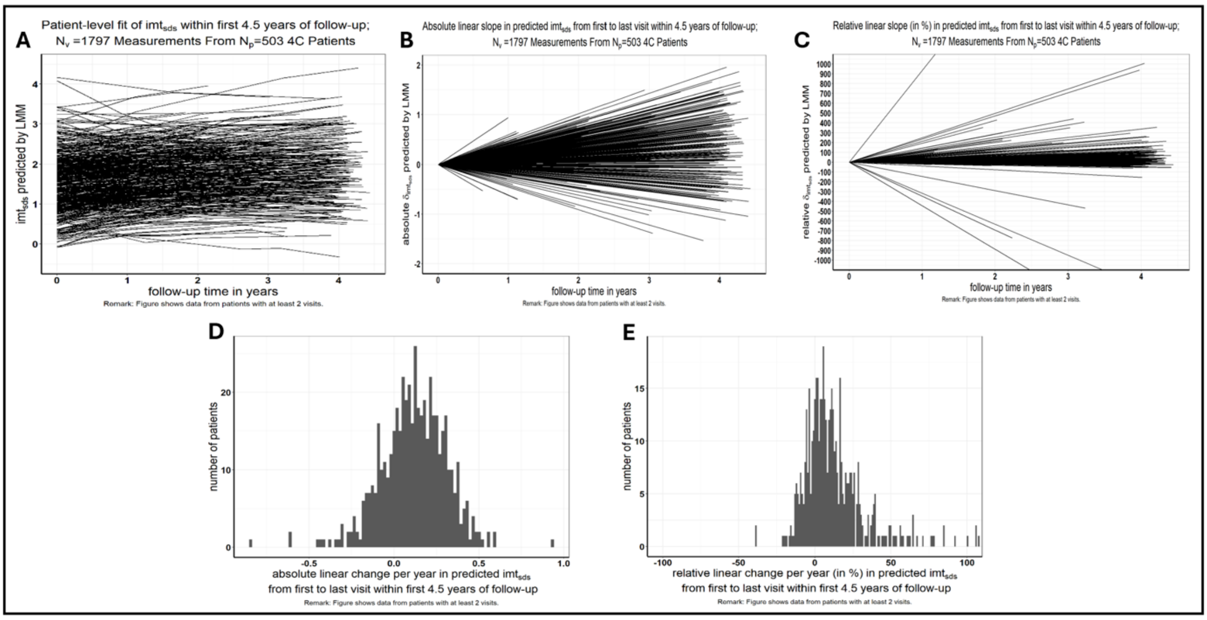


Figure S4: Derivation of absolute and relative change per year in cIMT SDS

**Patient-level absolute change per year in systolic blood pressure SDS**

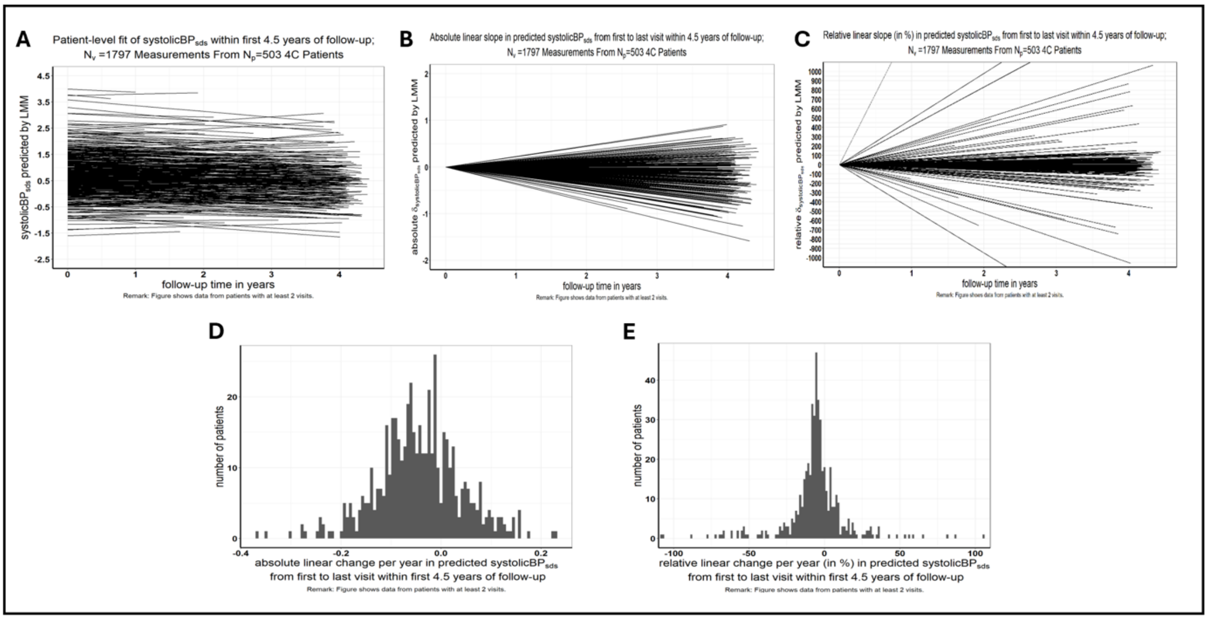


Figure S5: Derivation of absolute and relative change per year in systolic BP SDS

**Patient-level absolute changer per year in diastolic blood pressure SDS**

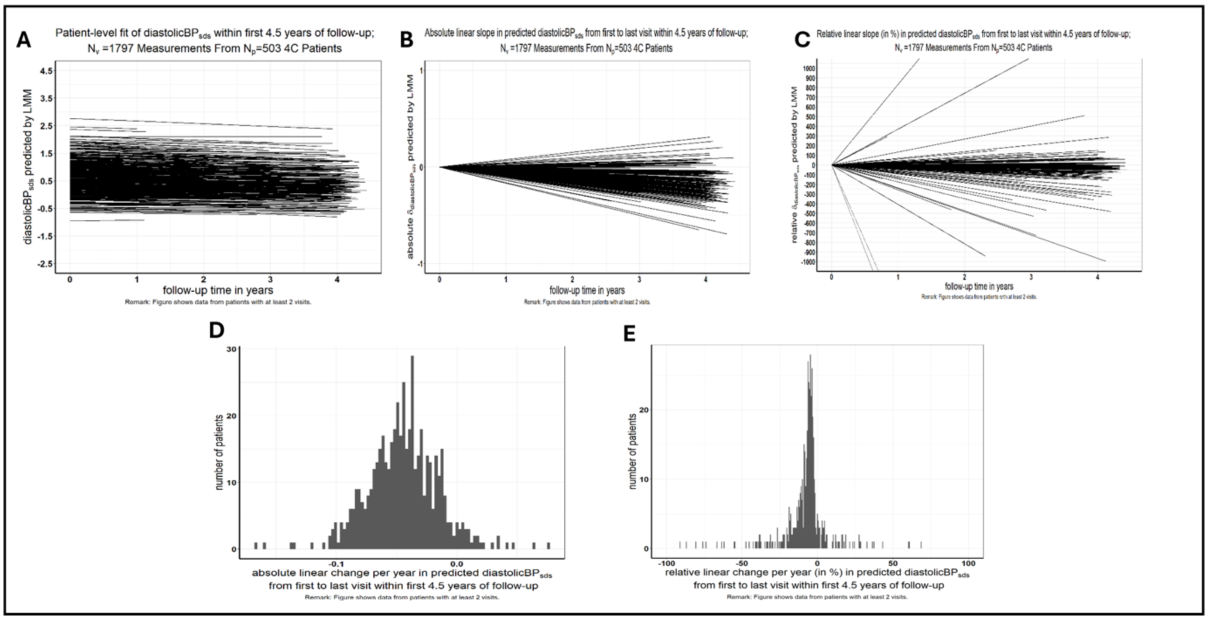


Figure S6 Derivation of absolute and relative change per year in diastolic BP SDS

References of supplementary material, listing according to citations in main manuscript


12. Portet S. A primer on model selection using the Akaike Information Criterion. *Infectious Disease Modelling*. 2020;5:111–128.

13. Rubin DB. *Multiple Imputation for Nonresponse in Surveys*. John Wiley & Sons; 2004.

15. R Core Team. R: A language and environment for statistical computing. R Foundation for Statistical Computing, Vienna, Austria. 2021.

16. RStudio R. RStudio: Integrated Development for R. http://www.rstudio.com/ RStudio, PBC, Boston, MA. 2020.

49. Little RJ, Rubin DB. *Statistical Analysis with Missing Data, Third Edition*. John Wiley and Sons; 2019.

50. Buuren S van, Groothuis-Oudshoorn K. *MICE: Multivariate Imputation by Chained Equations in R*. University of California, Los Angeles; 2010.

51. Robitzsch A, Grund S. Multiple Imputation Functions, Especially for “mice”. R package version 3.16-18. 2023.

52. Kleinke K. Multiple Imputation Under Violated Distributional Assumptions: A Systematic Evaluation of the Assumed Robustness of Predictive Mean Matching. *Journal of Educational and Behavioral Statistics*. 2016;42(4):371–404.

53. Edwards LJ, Muller KE, Wolfinger RD, Qaqish BF, Schabenberger O. An R2 statistic for fixed effects in the linear mixed model. *Statistics in Medicine*. 2008;27(29):6137–6157.
